# A disease model for Diffuse Intrinsic Pontine Glioma (DIPG) with mutations in TP53 and its application for drug repurposing

**DOI:** 10.1101/2022.06.22.22276788

**Authors:** Anton Yuryev, Anastasia Nesterova, Sergey Sozhin, Maria Shkrob

**Affiliations:** Elsevier

## Abstract

Brain cancers are ones of most aggressive and difficult to treat cancers. Despite numerous studies of the cellular mechanisms of gliomas, it is difficult to stop tumor growth. A complex genetic and epigenetic nature of many gliomas and poorly known pathways of human neuron precursors maturation suggest turning to big data analysis to find new insights and directions for drug development. We developed in silico molecular models and predicted molecular switches in signaling cascades that maintain multipotency of neuronal precursor cells in diffuse intrinsic pontine glioma (DIPG) driven by the H3K27M mutation and mutations in the TP53 gene. Oncogenes and biomarkers were predicted based on transcriptomics and mutational genomics data from a cohort of 30 patients with DIPG analyzed using Elsevier artificial intelligence methods and a collection of manually curated cancer hallmark pathways. The molecular models of DIPG with mutations in TP53 and histone 3 gene describe the mechanism of oligodendrocyte dedifferentiation due to activation of transcriptional factors OLIG2, SOX2 and POU5F1, epithelial-to-mesenchymal transition via strong EGFR and TGFR signaling, enhanced cell response to hypoxia via HIF1A signaling, and enhanced angiogenesis by VEGFA overexpression. Using in silico analysis, we identified drugs capable of inhibiting mutant TP53: vorinostat, cisplatin, paclitaxel, and statins were top ranked drugs. The predicted drugs and oncogenes had individual patient-level differences that can be visualized with created DIPG model and may be useful for future research in the field of personalized medicine.

## 1.2 Introduction

Diffuse intrinsic pontine glioma (DIPG) is a pediatric brain cancer diagnosed in 150-300 children in the USA per year (dipg.org). DIPG was previously diagnosed based on anatomical origin of the tumor in the developing midline brain pons. About 80% of anatomically defined DIPG tumors have somatic mutations in histone 3 genes where substitution of lysine 27 to methionine causes loss of trimethylation at that position (H3K27me3) (Lowe et al., 2019). Most often affected histone 3 gene is H3F3A (H3.3A) (Khuong-Quang et al., 2012), in rare cases mutations affect HIST1H3B or HIST1H3C (H3.1). Age of diagnosis, phenotype, location, and prognosis of DIPG depends on the isoform of histone 3 involved. The mean age of diagnosis of H3.3A tumors is 9.4 years, they have oligodendroglial-like phenotype, are found in the pons and other midline locations, and are more aggressive, with overall survival of 9 months. Mean age of diagnosis for patients with H3.1 mutant tumors is 5.9 years. H3.1 tumors usually have mesenchymal-like phenotype, are restricted to the pons, and have an overall survival of 15 months (Castel et al., 2015; Chen et al., 2021; Filbin et al., 2018; Loveson and Fillmore, 2018). Based on these observations, it was recently proposed to classify pediatric tumors based on their mutational profiles (Louis et al., 2016; Mosaab et al., 2020).

Studies of animal models suggested that growth of DIPG with H3.1K27M or H3.3K27M may require synergistic mutations in additional genes, such as ATRX, TP53, ACVR1 and PIK3CA (Fortin et al., 2020).

We used whole genome sequencing (WGS) and RNA-seq samples of DIPG patients from Open Pediatric Brain Tumor Atlas (OpenPBTA, Shapiro et al., 2021) to investigate the link between patients’ mutational profiles and cancer driving molecular mechanisms and to suggest drug candidates for different DIPG types. We selected for our analysis patients with mutations in TP53 and mutations in histone 3. Several publications reported high rates of single variant polymorphisms (SNVs) in TP53 in DIPG samples, however, data is still insufficient to discuss establish the frequency of TP53 mutations in the DIPG in general. In 1993 Zhang reported that 8 from out of 13 pontine gliomas samples had missense point mutations in TP53. Buczkowicz and colleagues defined that TP53 had somatic mutations in 42–71% of 39 DIPG samples (Buczkowicz and Hawkins, 2015; Khuong-Quang et al., 2012). Pollack and colleagues reported that TP53 was mutated in most of 100 samples of high-grade astrocytoma and glioblastomas (Pollack et al., 2002). Finally, Werbrouck found that TP53 was mutated in 48% (35 from 72 persons) DIPG patients (Werbrouck et al., 2019). We could find 23 samples with mutations in histone 3 and at least one mutation in TP53 from OpenPBTA.

TP53 is a well-studied transcription factor which controls expression of several hundreds of genes. TP53 is a cancer suppressor, and loss of its function facilitates cancer growth. TP53 gene is one of the most frequently mutated genes in cancer cells. The effects of mutations on TP53 function mainly depend on the origin of cancer cells and tissue context. TP53 targets are mainly involved in the regulation of apoptosis, cell proliferation, and DNA repair.

The precise influence of different mutant TP53 proteins on cancer cells is under investigation, but in general TP53 missense mutations can facilitate cancer growth and be “gain-of function” mutations (Soussi et al., 2014). While the particular effect of mutations in TP53 maybe not specified enough due to different approaches on annotation of TP53 isoforms (Soussi et al., 2014), it is believed that most mutant TP53 proteins have missense SNVs in DNA-binding domain (DBD domain) (exons 5-8) which obstruct the recognition of canonical p53RE element by other proteins and even make mutant TP53 protein more stable in cytoplasm than wild TP53 (Ferraiuolo et al., 2017).

In addition, mutated TP53 may act as an onco-driver because it causes specific transcriptional and non-transcriptional changes in cell protein landscape, such as expression of noncanonical TP53 targets and suppression of canonical TP53 targets. Many missense TP53 mutants form hybrid complexes with multiple binding partners like TP63 (TP73, vitamin D nuclear receptor or others) resulting in altering of their transcriptional activity (Yamamoto and Iwakuma, 2018) and thus affecting cell proliferation.

Non-transcriptional effects of TP53 mutations were linked to the accumulation of aggregates of wild and mutant TP53 proteins. Some TP53 regulators (upstream signaling and posttranslational modifications) stabilize the mutant TP53 protein better than the wild TP53, which is still transcribed from the second allele in cancer cells. This results in accumulation of mutant TP53 and blocking of wild-type TP53 functions by forming inactive tetramers with wild-type TP53 proteins or by facilitating proteasome-dependent degradation of TP53 (Ferraiuolo et al., 2017). For example, BAG2/5 proteins stabilize mutant TP53 and protect it from lysosome degradation (Yue et al., 2017). However, many researchers highlight that mechanism of accumulation of mutant TP53 or degradation of wild TP53 depends on cellular context and type of mutation - amino-acid variation. Further, aggregates of wild and mutant TP53 proteins may cause misfolded protein stress leading to defects in proteolysis, protein oxidation, ubiquitin–proteasome system (UPS), lipid metabolism, mitochondria, and vesicle trafficking processes such as autophagy. For example, mutant TP53-depended control of endosome trafficking and recycling lead to increase in EGFR plasma membrane concentration and to integrins overactivation without any gene transcription regulation (Comel et al., 2014; Onodera et al., 2013).

We used artificial intelligence (AI) methods and seq samples of DIPG patients to create in silico molecular pathway models and predict cellular and molecular effects of mutated TP53 and H3.3K27M on tumor progression in analyzed DIPG samples. Although we could not find significant correlation between genes with mutations and their direct differential gene expression in DIPG samples, we identified which cell pathways, receptors and transcriptional factors may facilitate the tumor growth in DIPG. Finally, we used the list of major molecular regulators of DIPG pathways and predict which drugs could be repurposed to treat the disease for the disease based on a text-mining method and a network of literature

## 1.3 Results & Discussion

### 1.3.1 Frequency of somatic mutations in DIPG patients

Whole genome sequencing (WGS) analysis identified 9,296 genes with mutations in 32 brain tumor samples available from 30 DIPG patients from OpenPBTA (summary list is provided in Supplemental materials, file “PNOC003 gene mutation frequency”).

We analyzed mutations in exons of genes in DIPG tumors. H3F3A and TP53 were the only genes where mutations in exons were present in more than a half of the patients. 80% of the patients had mutations in H3F3A, 76% of the patients had mutations in TP53. 56% of the patients had both mutations in H3F3A and mutations in TP53.

There were several known missense mutations in TP53 gene in samples from OpenPBTA. Three missense mutations (rs11540652 (c.743G>A), rs730882000 (c.475G>C) and c.716A>G) appeared in two patients. Two novel SNVs previously not reported in dbSNP were found (Figure A, supplemental file “PNOC003 TP53 mutations”). In cancers the majority of missense point mutations in TP53 cause formation of a faulty protein and in 10% of cases loss of TP53 protein function (Baugh et al., 2018). There are known SNVs in TP53 gene that are called “hotspots” because they are often reported in cancer cells. Hotspots commonly occur in TP53 DNA-binding domain between residues 102–292 (393 amino acids in the full-length protein) (Baugh et al., 2018). 23% of analyzed patients had hotspot missense mutations in exons 5, 7 and 8 of TP53 gene.

**Figure A.**
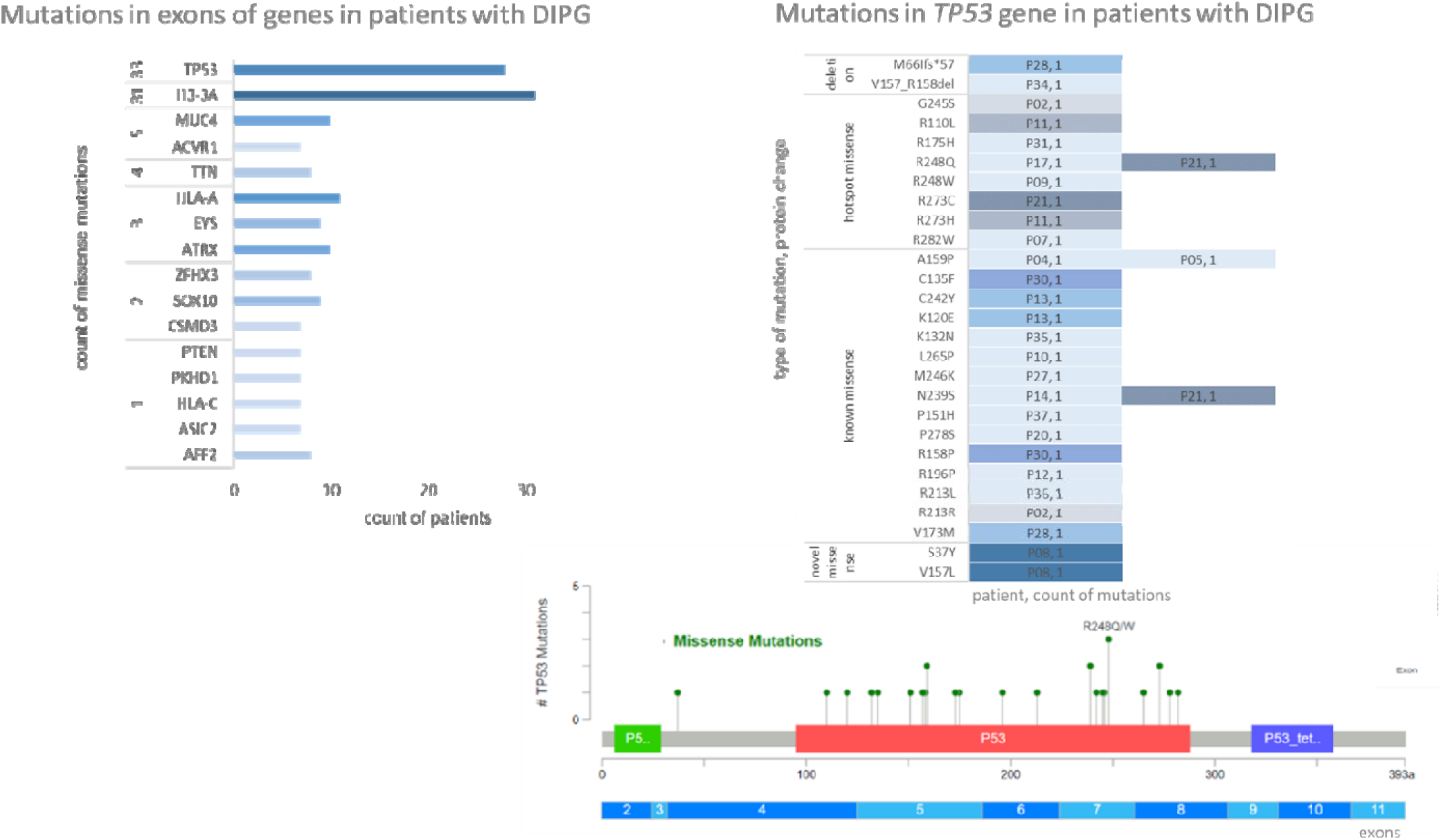
Genes with highest number of single polymorphism variants (SNVs) in brain tumor samples from 30 DIPG patients. Novel SNVs in TP53 are mutations that were not present in dbSNP database at the moment of research.

We identified additional 24 genes most frequently mutated in exon among DIPG patients. They were selected as 70 percentiles of all genes with mutations in openPBTA cohort. This list includes genes previously linked to DIPG (ACVR1 and ATRX) and genes known to be frequently mutated in other cancers (Figure A, Supplemental file “PNOC003 mutations frequency”).

We found missense exons mutations in MUC4 and ATRX in 30% of OpenPBTA DIPG samples. These genes were among top 10 mutated genes in all types of pediatric gliomas based on Wang et. al study. In glioblastoma UC4 was found overexpressed and may regulate invasion of cancer cells via EGFR signaling (Li et al., 2014). Muta ions in ATRX in H3.3A dependent DIPG (present in 5–12.5% cases) were described as the secondary event mutations (Haase et al., 2018). H3.3K27M along with mutations in ATRX and TP53 caused glioma tumors in vivo, and additional overexpression of PDGFRA lead to even more aggressive tumor invasion (Pathania et al., 2017).

ACVR1 gene has exon missense mutations in 23% of OpentPBTA DIPG samples. Activation mutations in ACVR1 reported in 20-32% of studied cases result in increased BMP signaling and are clinically significant for DIPG (Kluiver, 2020). Knockout of H3.1 and ACVR1 in Oligo2+ oligodendrocyte precursor cells (OPCs) resulted in high proliferation and arrest of differentiation of glia but did not lead to development of tumor cell phenotype.

HLA genes (HLA-A/B/C, HLA-DRA) had missense exon mutations in 33% of the analyzed tumors (10 patients). The HLA-A gene is necessary for activation of cytotoxic T-cells and T-helper-1-cell-mediated immune response against cancer cells with H3K27M mutations (Ochs K, 2017). HLA-B*15:17, HLA-A*68:02, HLA-A*02:05, HLA- C*12:03, or HLA-C*03:04 alleles were predicted to present peptides containing H3K27M mutations (Chang TC, 2017). Thus, the alterations of the immune response through impairment of HLA function may be important for H3.3A dependent DIPG development.

Several most frequently mutated genes in analyzed DIPG samples, such as titin (TTN), PIK3CA, were reported to be highly mutated genes in other cancers and were suggested as biomarkers of “tumor mutational burden” the condition of hypermutated tumors due to genomic instability (Oh et al., 2020). On the other hand, it was reported that spontaneous high-grade gliomas only developed if PIK3CA gene had gain-of-function mutation in combination with mutations in Hist1h3b and ACVR1 (Fortin et al., 2020).

WGS data from OpenPBTA revealed high mutation rate in 5’-UTR intron of protein receptor phosphatase PTPRD gene (present in 25 patients) (Figure A, Supplemental file “PNOC003 mutations frequency”). Additional 4 DIPG patients had mutations in other PTPRD introns. Most cancer mutations studies use Whole Exome Sequencing (WES) and transcriptomics (RNAseq) data and therefore focus on mutations in exons which are easier to interpret. Mutations in PTPRD were not localized in any evolutionary conserved gene region. PTPRD is the third longest gene (230Mb) in the human genome, so these mutations may have accumulated at random during tumor evolution. But interestingly, the number of somatic mutations in other long genes (RBFOX1 (248 Mb), CNTNAP2 (230 Mb), DMD (220 Mb), DLG2 (218 Mb), MACROD2 (206 Mb), EYS (199 Mb)) in DIPG patients was much lower than in PTPRD. The next highest number of intron mutations was found in EYS gene: 28 mutations in introns in 9 patients. It is not clear how point mutations in introns of PTPRD gene affect the function of the protein and whether they are clinically significant. Nonfunctional PTPRD have been reported in other gliomas (Veeriah et al., 2009) and its loss was found to cause aberrant STAT3 activation promoting glioma (Ortiz et al., 2014). In adult brains PTPRD is involved in trans-synaptic interaction between neurons (Yoshida et al., 2011) and mediates cortical basal dendritic arborization, synaptic plasticity and neuronal growth in perineuronal nets (Carulli and Verhaagen, 2021; Cornejo et al., 2021).

In general our results are consistent with frequencies of clinically significant mutations in DIPG reported in previous studies (Argersinger, et al., 2021; Buczkowicz and Hawkins, 2015).

### 1.3.2 Prediction of oncogenes and tumor suppressors in TP53-dependent DIPG cancer growth

We present methodology of “pathway activity analysis” which can predict cancer driving molecular mechanisms based on the analysis of patient transcriptomics and genomics data using gene regulatory network compiled from independent observations reported in peer-reviewed scientific literature.

“Pathway activity analysis” can be used in addition to the analysis of mutation frequency to predict gene’s impact on DIPG tumor growth and to suggest oncogenes and tumor suppressors. We define oncogenes and tumor suppressors as either genes with mutations or key modulators of intracellular signaling downstream of mutated genes which contribute (oncogenes) or prevent (tumor suppressors) tumor growth.

We tested the predictability of oncogenes and tumor suppresses in individual patient tumor by cross-matching mutated genes with statistically significant regulators of differentially expressed genes and cancer pathways.

As a starting point we used RNA-seq data of 34 DIPG tumor samples with TP53 and H3.3A mutations from OpenPBTA to determine differently expressed (DE) genes (Supplemental file “PNOC003vsGSE120046”). Single cell transcriptome of embryonic pons was chosen as control for calculating differential expression in tumor samples. To find the best healthy tissue control for gene expression analysis we have considered three public datasets measuring normal brain tissue (Fan et al., n.d.; Lindsay et al., 2016; Love et al., 2014). Fan et al, 2020 dataset with measuring single cells transcriptome of embryonic pons appeared to be the best dataset for our control selection criteria and therefore it was selected for analysis of all patients (see Methods).

#### 1.3.2.1 Prediction of “gene expression regulators”

The first step in “pathway activity analysis” is to define a list of statistically significant protein regulators of DE genes (“gene expression regulators”) based on the literature-extracted information in Elsevier Biology Knowledge Graph 2020, which covers direct and indirect effects of proteins on the expression of target genes at mRNA or protein level (see Methods). Importantly, this approach helps to avoid missing players that change their activity but not mRNA levels, which is a common situation, since there is only a 40% correlation between protein activity and levels of mRNA expression in vivo (Vogel 2012). Determining “gene expression regulators” from literature-derived gene regulatory network networks which has evidence about all levels of expression helps us to avoid missing regulators that are not detected on mRNA expression level.

To find “gene expression regulators” we looked for proteins which are: 1) upstream regulators of a subset of DE genes in DIPG tumors; 2) regulate targets with DE above statistically significant threshold in a concordant manner. “Gene expression regulators” were ranked by their “activity score”, calculated using Sub-Network Enrichment Analysis (SNEA) (see Methods). Positive activity score indicates that “gene expression regulator” induces the expression of upregulated targets, negative activity score indicates that “gene expression regulator” induces predominantly down-expressed targets. Regulators with positive activity score henceforth we will define as “regulators of active sub-networks or “active regulators”. Regulators with negative scores as “regulators of suppressed sub-networks”.

In all 34 samples (from 32 patients) we found 2,335 “gene expression regulators” with SNEA p-values less than 0.05 in at least one of the samples from OpenPBTA. We then excluded samples without TP53 mutations and two outliers (P08, P014) (Supplemental Figure 1). The final list contained 926 regulators that were statistically significant in at least one of 21 tumors which had mutations both in histone 3 and TP53 (Figure B, Supplementary file “Regulators of gene expression”).

**Figure B.**
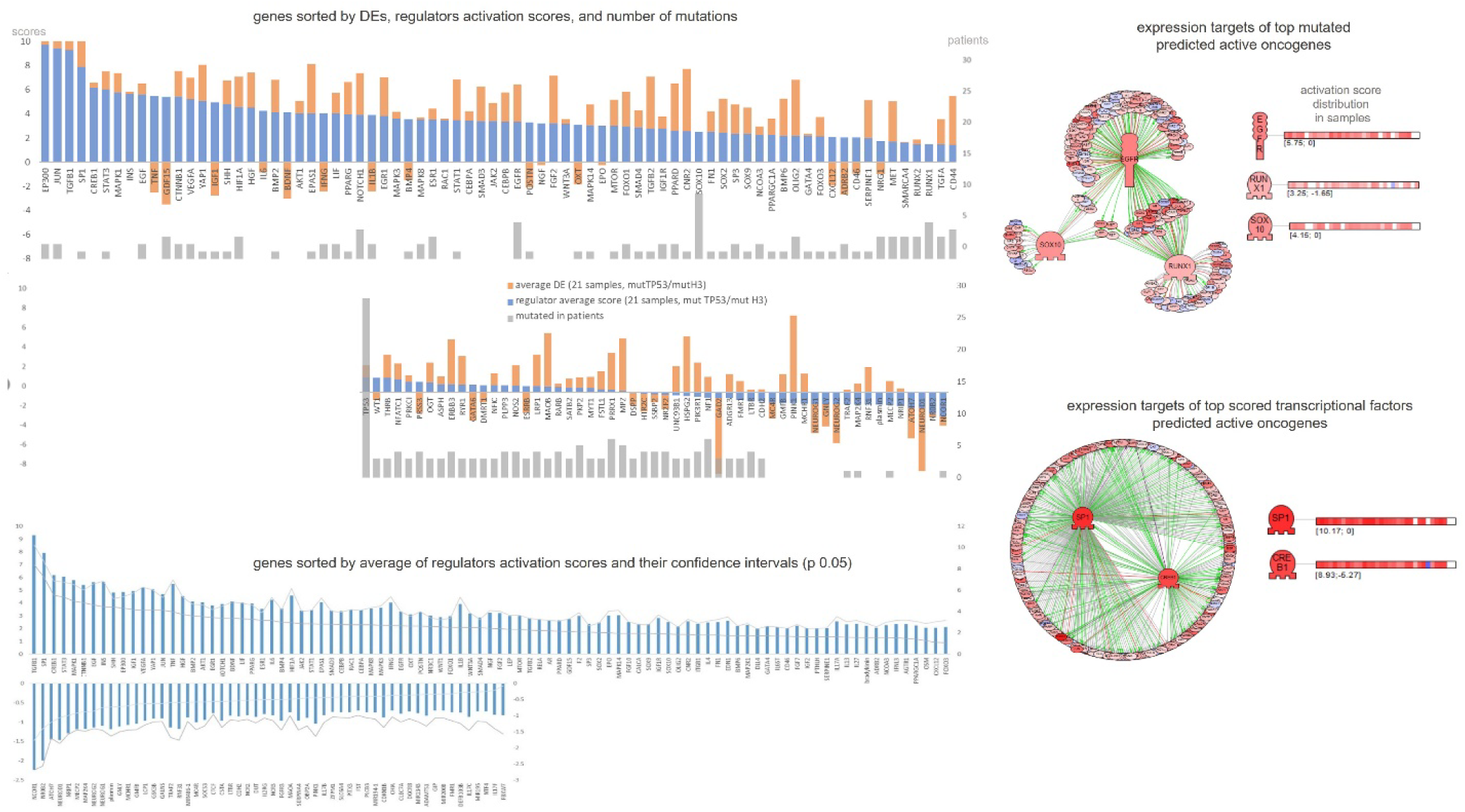
Statistically significant gene expression regulators based on the analysis of 21 DIPG patients with mutations in TP53 and histone 3. Regulators with activation scores above 2 and below -1 are shown. Sub- networks are examples of active EGFR, RUNX1, SOX10, SP1 and CREB1 regulators and their expressional targets in Elsevier Knowledge Graph. See more data in supplemental materials, file “4_Regulators of gene expression”.

Top three active regulators in this list were receptor TGFBR2, transcription factors SP1 and CREB1 (Figure B). In accordance with literature we also found EGF/EGFR and PDGFRA were statistically significant active regulators. EGFR was a significant regulator in 90% of samples, while PDGFRA in 52%. Previous studies showed that an extra-amplification status of platelet-derived growth factor receptors (PDGFRs) and EGFR in DIPG may drive tumor growth in DIPG (Kluiver et al., 2020; Paugh et al., 2011; Puget et al., 2012).

Transcription factors NCOR1, NROB2 and NEUROD1 had the smallest average negative activation score (in 90%, 80% and 85% patients respectively). Expression levels of NCOR1 and NEUROD1 genes themselves were downregulated in the majority of DIPG tumors (31 and 22 out of 32 samples).

Activity of gene expression regulators did not correlate with their mutation status in analyzed patient cohort (Figure B) with exception of TP53. Only three high-score regulators (SOX10, EGFR, and RUNX1) had mutations in several patients. It may suggest that mutations in these genes contribute to cancer growth. Below, we analyzed the interactions between found regulators to explore their role as onco-drivers in DIPG.

#### 1.3.2.2 Cells in DIPG tumor

Meeting our expectations, SNEA analysis showed that DIPG samples were enriched with markers of cells from brain pons and embryonal pons (see Methods). We found the following cell types enriched with gene expression regulators in DIPG: neuron, oligodendrocyte precursor cell, stem cell, astrocyte, embryonic, cancer cell lines, vascular epithelial cells, T-cells including NK-cytotoxic cells, and tumor infiltrating macrophages (Supplementary file “Cells enriched with regulators”).

Neural stem cells (NSCs) give rise to neurons, astrocytes, and oligodendrocyte precursor cells (OPCs) which can in turn differentiate into oligodendrocytes. Average time of DIPG diagnosis (6 years old) corresponds to normal physiological expansion of neuron migration, myelination and OPCs differentiation in human pons. Healthy pre-OPCs are known to express markers OLIG2 and NEUROG2, while mature OPCs may have high levels of PDGFRA, OLIG2, NKX2.2, NKX6.2, SOX10 and NG2 (CSPG4) expression. In 80% of DIPG cases pluripotent OPCs were shown to express nestin, vimentin and the same markers as healthy OPCs (SHH-responding OLIG2, PDGFRA, NG2, OLIG1 and OLIG2) (Loveson and Fillmore, 2018; Swartling et al., 2014).

In analyzed DIPG samples several OPCs markers (SOXs, OLIGs, nestin, CSPG4, etc.) were simultaneously overexpressed and came out as active regulators (Supplementary Figure 3). SOX10 and OLIG2 had mutations in 9 and 3 patients respectively (Figure B). Markers for other neural cells such as astrocyte (SOX9, GFAP, etc.), as well as microglia, and endothelial vascular cells were also found to be active regulators in DIPG tumor samples by our analysis (Supplementary Figure 3).

Thus, we supported previously published conclusions that DIPG tumor is a heterogenous tumor originating mainly from the lineage of oligodendrocytes (primitive oligodendrocyte precursor cells (OPC)-like progenitor (pri-OPCs/pre-OPCs) population) (Filbin et al., 2018; Loveson and Fillmore, 2018; Venteicher et al., 2017; Weng et al., 2019). Further, our analysis supports the hypothesis that somatic DIPG driver mutations in histone 3 and other genes occur in progenitors of neuronal stem cells - OPCs (Weng et al., 2019; Zong et al., 2015) arresting their further differentiation. However, the possibility of additional mutations (e.g., PDGFR amplifications) in astrocytes or other neuronal lineage cells is also discussed (Jones et al., 2017; Phillips et al., 2013). Somatic mutations in non-neuronal cell lineage (microglial-mesodermal, vascular-mesenchymal, immune cells) are not discussed in the literature as the drivers of the DIPG tumor cells microenvironment.

#### 1.3.2.3 Prediction of cancer hallmarks and molecular mechanisms (pathways) of TP53- dependent DIPG cancer growth

To suggest that an active gene expression regulator is an onco-driver, it is important to understand the molecular mechanism of its contribution to tumor growth. We conducted “pathway activity analysis” to link 926 statistically significant “gene expression regulators” identified at the previous step to cell processes and signaling cascades contributing to or affected by DIPG tumor growth. Pathways enriched with active “gene expression regulators” were determined using Gene Set Enrichment Analysis (GSEA) and “average activity” scores of regulators across 21 DIPG patients were calculated. Statistical significance of pathway activation was determined by GSEA p-value (see Methods).

Results of enrichment analysis highly depend on the input collections of pathways or protein groups (Nesterova, et al., 2020). For example, analysis using Gene Ontology, the most popular collection for enrichment algorithms, returned very broad and ambiguous cell processes, such as “regulation of cell proliferation” and “DNA-binding” (Supplementary file “Pathways enriched with activity regulators”). More specific and relevant results were generated using Elsevier’s Pathway Collection, which contains 3100 pathways, including 225 cancer models organized by 14 cancer hallmarks (Table 1, supplementary file “Pathways enriched with activity regulators”), including ten originally proposed hallmarks (Hanahan and Weinberg, 2011) and four additional hallmarks: mitochondrial instability, tumor microenvironment, drug resistance and cancer epigenetics.

**Table 1.** Top ranked pathways and respective cancer hallmarks enriched with regulators of gene expression in DIPG tumors with mutations in TP53 and histone 3.

| Pathway | Cancer hallmark | Pathway score |
| --- | --- | --- |
| VEGFA Dependent Angiogenesis in Cancer | Inducing Angiogenesis | 21.4 |
| Epithelial to Mesenchymal Transition in Cancer | Activating Invasion and Metastasis | 21.2 |
| Glioma Stem Cell Program Activation | Sustaining Proliferative Signaling | 20.8 |
| Growth Factors Signaling in Neuroblastoma | Resisting Cell Death | 16.1 |
| STAT3 and NFkB Activate Inflammation-Induced Tumorigenesis | Tumor-Promoting Inflammation | 15.6 |
| Cancer Cells Inhibit Adipocyte Differentiation | Deregulated Metabolism | 13.2 |
| Hyaluronic Acid, CD44 and HMMR in Cancer Cells Motility, Invasion, Proliferation and Survival | Tumor Microenvironment Physics Adaptation | 12.7 |
| Hedgehog Signaling Activation by Blocking of Tumor Suppressors | Evading Growth Suppressors | 12.3 |
| IDO1 in Cancer Immune Escape | Evading Immune Destruction | 12.3 |
| TERT Activation in Cancer | Enabling Replicative Immortality | 10.7 |
| Acetylases Inhibition in Histone Deacetylation in Cancer | Genome Instability | 9.6 |
| Possible Interplay between SOX11, CCND1 and EZH2 in Mantle Cell Lymphoma | Cancer Epigenetics | 6.6 |

Four major cancer hallmarks enriched with “gene expression regulators” are “Sustaining Proliferative Signaling” (22 pathways with maximum ranks), “Activating Invasion and Metastasis” (21 pathways), “Inducing Angiogenesis” (15 pathways) and “Tumor Promoting Inflammation” (16 pathways). Accordingly, top ranked pathways describe cell behavior during angiogenesis in cancer, epithelial to mesenchymal transduction and tumor-related inflammation (Table 1, Figure C, supplementary file “Pathways enriched with activity regulators”). In the next section we discuss highest-scoring pathways and possible roles of mutated genes and regulators of gene expression as oncogenes and onco-suppressors in analyzed DIPG tumors.

**Figure C.**
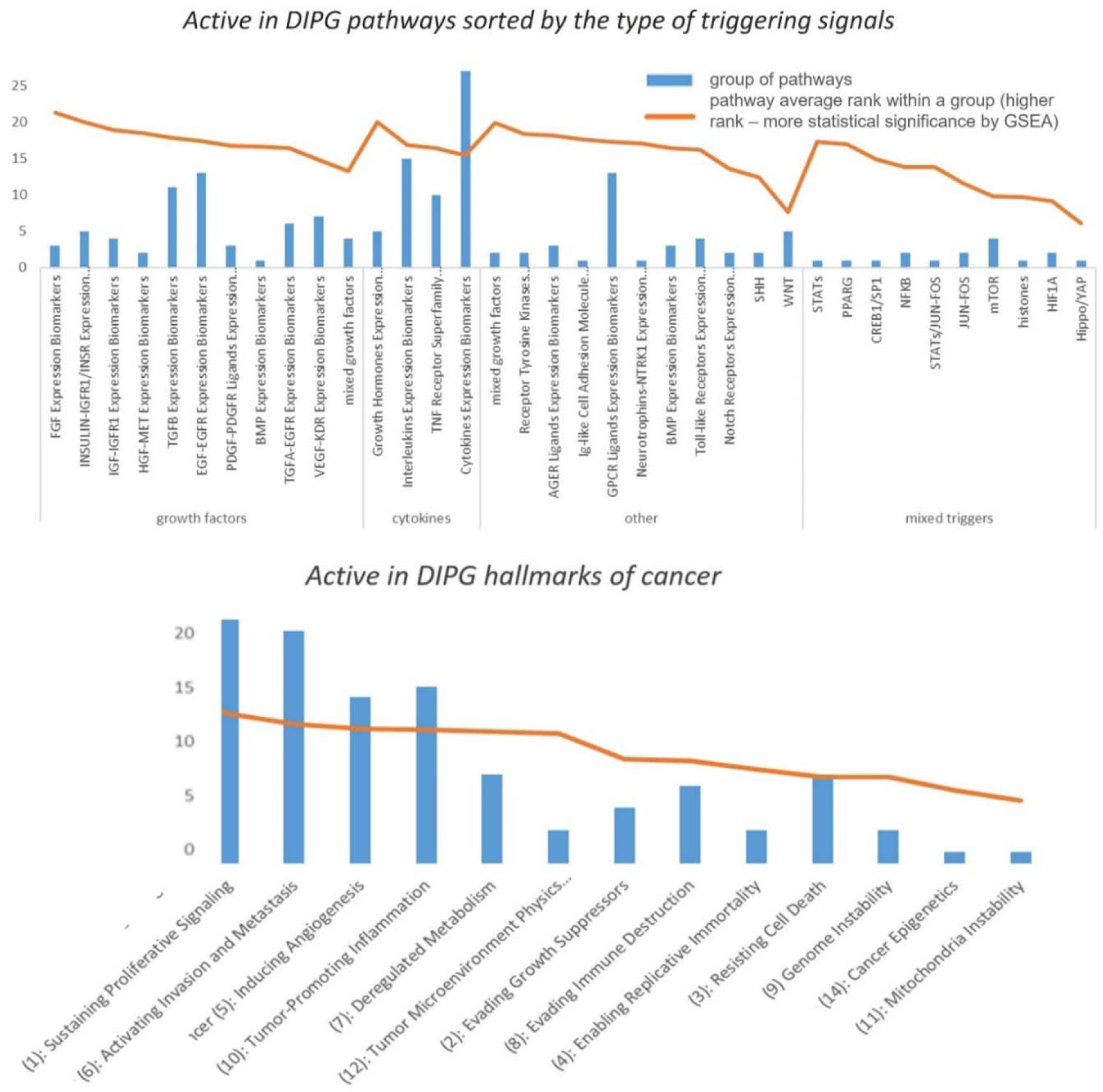
Statistically significant pathways from Elsevier Pathway Collection (including cancer hallmarks) sorted by relevant for DIPG cells processes, types of major transcription factors or receptor triggering signals. Pathways can be opened as web links (see supplemental materials).

### 1.3.3 Pathway activity analysis for predicted oncogenes and tumor suppressors in DIPG

Active signal transduction pathways in DIPG identified by pathway analysis are consistent with previously published distinctive mechanisms guiding cancer cell growth (Duchatel, et al., 2019; Puget et al., 2012). To present results of “pathway activity analysis” as a single simplified cell signal transduction pathway we combined top ranked signaling molecules such as growth factors (especially EGF, FGF, PDGF and TGFb family) and cytokines (interleukins, TNF). We also included in the model top ranked canonical embryonal cascades NOTCH, SHH, Hippo/YAP and WNT which maintain proliferation and pluripotency of cancer cell. As a result, combined top 87 active regulators, 23 “regulators of suppressed sub-networks” (ones with top negative scores) and their interactions from cancer hallmark pathways shaped the model “The pathways of DIPG regulators” (Supplemental Figures 2 and 2.1). In the model, ligands pass signals into the tumor cell through the respective receptors to activate calcium, PIP3-AKT, MAPKs and cAMP cascades which then triggers interplay between transcription factors (SMADs, JUN-FOS, STAT3, HIF1A, CREB1 and others). These transcription factors are well known oncogenes that further induce expression of many proteins including initial extracellular triggers – growth factors, cytokines, and their receptors. Such signal amplification through transcriptional positive feedback loop may fuel uncontrollable growth of tumor cells.

#### 1.3.3.1 Sustaining Proliferative Signals Hallmark

Proliferation pathways were among the top results of “pathway activity analysis” because they were enriched with overexpressed members of AKT1, mTOR, MAPKKs, calcium cascades and related transcription factors that control expression of cell cycle proteins. Signals that initiate proliferation, such as EGF and EGFR1, growth factors (TGFB, IGF), SHH and WNT, were among top scored regulators in those pathways.

FGF, EGF, SHH and PDGF signaling cascades are considered to be the primary inductors of proliferation and differentiation of normal oligodendrocytes, the most proliferative cells in the brain (Lanjewar and Sloan, 2021). Signaling cascades via EGF group receptors (EGFR, ERBB3 and ERBB4) initiate proliferation of many cancer cell types, including glioma cells (Rajaram et al., 2017; Swartling et al., 2014). We found that 71% of classic EGFRs signaling components were over-expressed in DIPG samples (S. Figure 5).

PDGFR signaling tightly interacts with EGFR/ERBBs signaling and controls activation of MAPK, AKT and calcium cascades. PDGFR amplification is considered one of possible driver events for DIPG tumor growth (Hoeman, et al., 2018). We found that although both PDGFB and PDGFRA were among significant active regulators in DIPG samples, their average activation scores among 21 patients were not the highest (0.7 and 0.5 in interval -2.2 to 9) because their known expression targets were not highly over-expressed in DIPG samples. This observation may indicate that in analyzed samples, PDGFR receptor signaling is not hyperactivated.

FGF2 signaling (estimated by “FGF2 -> STAT Expression Targets”, “FGF2 -> AP-1/CREB/CREBBP/ELK/SRF/MYC Expression Targets”) had the highest average pathway activity score in our analysis. FGF2/FGFR3 signaling was shown to inhibit astrocytes differentiation and promote oligodendrocyte lineage. Expression of FGFR3 was linked to late OPCs progenitors that already enter to terminal differentiation stage, which is happening around 6-9 years in normal pons development (Inglis-Broadgate et al., 2005; Oh et al., 2003). Indeed, some transcription factors downstream of FGF receptors are effectively activate proliferation and were over-expressed in DIPG samples. However, most of expression targets of FGF2 and FGFR3 were downexpressed, and FGF2 was “regulator of suppressed sub-networks” (with negative activation score -0.97, in -2.2; 9 interval). FGF2 and FGFR3 themselves were only slightly over-expressed in analyzed DIPG tumor samples (see Supplemental file “PNOC003vsGSE120046”).

Based on our analysis, we conclude that EGFs and SHH but not PDGFRA and FGFs are likely major proliferative signals in analyzed DIPG tumors.

#### 1.3.3.2 Tumour Promoting Inflammation Hallmark

Previous findings demonstrate minimal immune activation in the DIPG microenvironment in contrast to other types of brain tumors. T-cells including NK-cells are not considered to be highly involved in DIPG infiltration. It was reported that proteins involved in T-cell promoting IL2 cascade had reduced expression in diffuse gliomas, but T-cell suppressing IL8 and TGFB cascades had typically have higher level of expression in diffuse gliomas (Lin et al., 2018; Price et al., 2021).

We also found that components of T-cell receptor complex, IL8, IL15, CD55, CD36 and their receptors were down-expressed in DIPG samples. These genes as well as HLA genes had been mutated in 3 or more patients. Other proteins important for immune cell function had top negative scores (“regulators of suppressed sub- networks”): receptors (IL2RG, IL4R, CD36, CD14), transcription factor GATA3 or UNC93B1 (TLR signaling regulator), granulysin (GNLY, secreted from cytotoxic T cells). Based on in silico analysis of 495 patients with pediatric gliomas, Wang with co-authors recently suggested to classify gliomas into 3 immune subtypes: “immune hot” (IS-I), “immune altered” (IS-II), and “immune cold” (IS-III). DIPG tumors were classified as IS-II and IS-III (Wang et al., 2021). Our transcriptomics analysis also found DIPG tumors to be noninflammatory with low antitumor immune activity.

However, results of “pathway activity analysis” indicated that inflammatory mediators TGFb, INFG, TLRs, STAT3, and NFkB were active gene expression regulators and were over-expressed in DIPG samples compared with embryonic cells from healthy pons. Such results may reflect the activity of microglia resident macrophages (tumor-associated macrophages (TAMs) (Low and Ginhoux, 2018).

TAMs heavily infiltrate pons in DIPG and form immune and inflammatory environment in a tumor (Price, et al., 2021; Ross et al., 2021). TAMs in DIPG were reported to have unique characteristics: they expressed CD45 (PTPRC), CD68, and CD163 which is partially mediated through PDGFB signaling (Price et al., 2021). The amplification of PDGFR in DIPG cancer cells (OPCs or astrocytes), astrocyte derived cholesterol and cytokines (especially IL34 and TGFB2) are known to promote microglial maturation and proliferation (Butovsky et al., 2014).

Microglia in healthy brain expresses numerous classical macrophage markers (CD11b, CSF1, CD15, CD172a, CX3CR1) and more specific EGR1, P2ry12, TMEM119 and Siglec-H (based on mice models) (Low and Ginhoux, 2018), TAM family receptors/ligands and their targets (Butovsky et al., 2014).

Not “tumor-associated macrophages” but “central nervous system macrophages” were the top ranked cell type according to our SNEA analysis (Suppl. Table 6_Cells enriched with regulators (922).xlsx) due to highly overexpressed markers IFNB1, TGFB1, SPI1, LRP1, APP, NR3C2, CCL2, and CSF1R.

In general, reported CD markers of TAMs in analyzed DIPG samples were down expressed. However, CD45 and EGR1 were statistically significant regulators of over-expressed genes. Also, two innate immunity receptors (TAM receptor protein tyrosine kinases AXL, TYRO3) were overexpressed in samples and can be expression targets for top scored regulators SP1, YAP1 and TP53 (S. Figure 4).

#### 1.3.3.3 Inducing Angiogenesis Hallmark

Cancer hallmark pathway “Inducing Angiogenesis” had the highest pathway activity score because corresponding pathways have been enriched with oxygen-driven transcription factors such as HIF1A, SP1 and STAT3 that cause synthesis of master inductors of angiogenesis (VEGFA, PDGF, FGF2) and their receptors. In addition, RUNX1, well known regulator of hematopoiesis, was a significant active regulator in 24 DIPG patients (1.49 score in interval [-2.2; 92]) and had mutations in 4 patients (1 patient with SNV in exon and 3 in introns).

Histologically, DIPG tumors have been reported to show no neovascularization and have a functional blood-brain barrier (BBB) (Plessier et al., 2017). In vivo models showed that vascular endothelial functions in DIPG did not differ from normal unlike pediatric high-grade glioma with tumor disorganized angiogenesis and blood–brain barrier dysfunction (Wei et al., 2021). Moreover, some researchers suggests that brain pons and pons tumors have superior BBB which strongly restricts the passage of substances into vessels (Loveson and Fillmore, 2018; Plessier et al., 2017; Wei et al., 2021).

We did not have the information regarding the histological profile of analyzed DIPG tumors, however, our results point to not transformed but more intense molecular signaling control of the vascularization and endothelial cell function in DIPG. Intensive microglial migration and expansion which depends on blood vessels and vascularization (VEGFR signaling, MCP1/MIPA, etc.) may drive the angiogenesis (Conway et al., 2014). However, we found that cascades of CCL2 and CD36 (hemoglobin sensor) which trigger the release of angiogenesis signals by macrophages and epithelial cells were down-expressed in most of patients.

#### 1.3.3.4 Activating Invasion and Metastasis Hallmark

“Activating Invasion and Metastasis” cancer hallmark had the top score in “pathway activity analysis” due to enrichment with regulators that control ECM remodeling and epithelial-to-mesenchymal transition (ETM). ECM remodeling pathways include top scored active regulators in DIPG samples TGFb and NOTCH-dependent transcription factors, and key masters of transcription regulation HIF1, STAT3 and NF-kB. TGFb family members (TGFb and BMP ligands) trigger both cancer promoting and suppressing pathways depending on cell type. In general, it is known that canonical TGFb signaling via SMADs may have protective role in the early stages of some cancers but in later stages and in brain cancers it more likely promotes ETM of OPCs and cell migration (Loveson and Fillmore, 2018).

In healthy brains expanding microglia regulates density of OPCs and their differentiation (myelinogenesis) by inducing apoptosis via release of HSP60 signal and involvement of TLR4/NFKB pathways in OPCs (Low and Ginhoux, 2018). MMP8/9 and HSP60 were among down-expressed DE genes in analyzed samples, which may indicate the disruption of microglial function of controlling OPCs density in DIPG.

During mesenchymal transition transcription factors stimulate synthesis of biomarkers of mesenchymal (highly invasive) status and metalloproteases that degrade matrix. Metalloproteins typically active in cancer and markers of mesenchymal status of the cell (N-cadherin, vimentin, MMP8/9) were not over-expressed in DIPG samples comparing with control. However, more specific neuronal metalloproteases MMP24/25, and metalloproteinases MMP14/MMP1616 linked to cancer were highly over-expressed in analyzed DIPG samples. As well as tenascin family of extracellular matrix glycoproteins which involved in neural development, oligodendrocytes and cancer (Chiovaro et al., 2015; Jones and Jones, 2000) Previously, Tenascin-C (TNC) overexpression was linked with the H3K27M mutation and cell invasion in DIPG (Qi et al., 2019). TNC was on the list of active regulators (0.68 score) and was overexpressed in analyzed DIPG samples (average 3.86 log DE in interval [-10;13]).

In general, it is not well known how mutations affect invasion of cancer cells in DIPG. Mutated TP53 caused changes in SNAI1 function which were linked to enhanced cell migration in DIPG (Kluiver, et al., 2020; Qin et al., 2017). All major cell proliferative cascades with MAPKs, PI3K/AKT, Rho and STATs may initiate neuronal mesenchymal transition because of their ability to activate SNAI1/2, ZEB1/2, and TWIST1. SNAIs, ZEBs or TWIST1 were not statistically significant regulators of DE genes in DIPG samples and only ZEB1 had missense mutation in one patient from our DIPG cohort.

Proteins involved in actin organizations and pathways of Tight-Junction Assembly (TGP2, LLGL1), Focal Junction (integrins), Axonal transport (synaptopodins) were over-expressed in analyzed samples (91 proteins total). Action organization nucleoproteins AHNAK and AHNAK2 were among top 10 mutated genes in pediatric gliomas according to Wang et. al study (Wang et al., 2021). In samples we analyzed AHNAK was mutated in P05 and AHNAK2 had mutations in 4 samples. We detected AHNAK mRNA was over-expressed (5.9 average log DE values, 6.87 and log DE in P05) in DIPG tumors with TP53 mutations. It was reported that AHNAK inhibited lung cancer cell stemness by reducing TP53 tumor-protecting effects. AHNAK-TP53 complex was regulated by ubiquitin ligase (UBE3C) in lung cancer cells. In addition to being a scaffold protein in multi-protein complexes in nucleus, ANHAK reported to be membrane protein of the blood–brain barrier and cell architecture (Gu et al., 2019). ANHAK activated TGFB/SMAD3 mediated epithelial-mesenchymal transition in melanoma model in response to PDFG and EGF signals. The high level of ANHAK expression was reported as a marker of invasive ability of several cancers (mesothelioma, larynx carcinoma) (Sohn et al., 2018).

### 1.3.4 Modeling effects of interaction between mutant *TP53* and *H3.3A* and their targets in DIPG

Several reviews published the molecular model of DIPG (Duchatel, et al., 2019; Kluiver et al., 2020; Nagaraja et al., 2019; Puget et al., 2012) and highlighted a complex genetic nature of DIPG origin.

At the age of 6 years, the epigenetic silencing of histone 3 (by trimethylation H3K27me3) in oligodendrocytes of brain pons becomes very intense (Egawa, et al., 2019; Loveson and Fillmore, 2018). It is hypothesized that this is not happening in DIPG tumors where the oligodendrocyte precursors become cancer cells because of the combination of mutations that cause hypomethylation of histone 3 with de novo amplifications of proliferative genes (such as in AVCR1, PDGFRA) or/and with point mutations in others (TP53, ATRX, PPM1D, etc.).

However, this understanding is not sufficient to find drugs to stop DIPG growth, and that is why it is important to continue testing new hypotheses about existence of additional molecular switches in cascades that regulate DIPG. For example, epigenetic regulation (loss of repressive methylation marks in chromatin-transcription complexes) of genes specific to OPCs is poorly studied. Below, we discussed proteins and mechanisms predicted by our analysis that may cause gain-of functions and proliferation of cancer cell.

#### 1.3.4.1 Oncogenes and onco-suppressors based on “pathway activity analysis”

15 genes mutated in more than one patient may be involved in DIPG tumor growth according to the model of interaction between predicted regulators and pathway activity analysis (Figure D). Most active regulators within the model should be considered oncogenes since they maintain cell proliferation and activation of gene expression. SP1, regulator with the highest activation score (7.9), is a known oncogene and transcription activator involved in epigenetic regulation (Vizcaíno et al., 2015). Other oncogenes within the model include a) transcription factors involved in histone acetylation, cell proliferation, migration, and oligodendrocyte dedifferentiation (SP1, CREB1, STAT3, EP300, etc.); b) growth factors and their receptors which may trigger cell growth and migration (TGFB1, EGF, VEGF, etc.); c) intracellular signaling initiators of MAPK cascade or protein translation (PI3K, AKT, PLCs, MTOR, etc.).

**Figure D.**
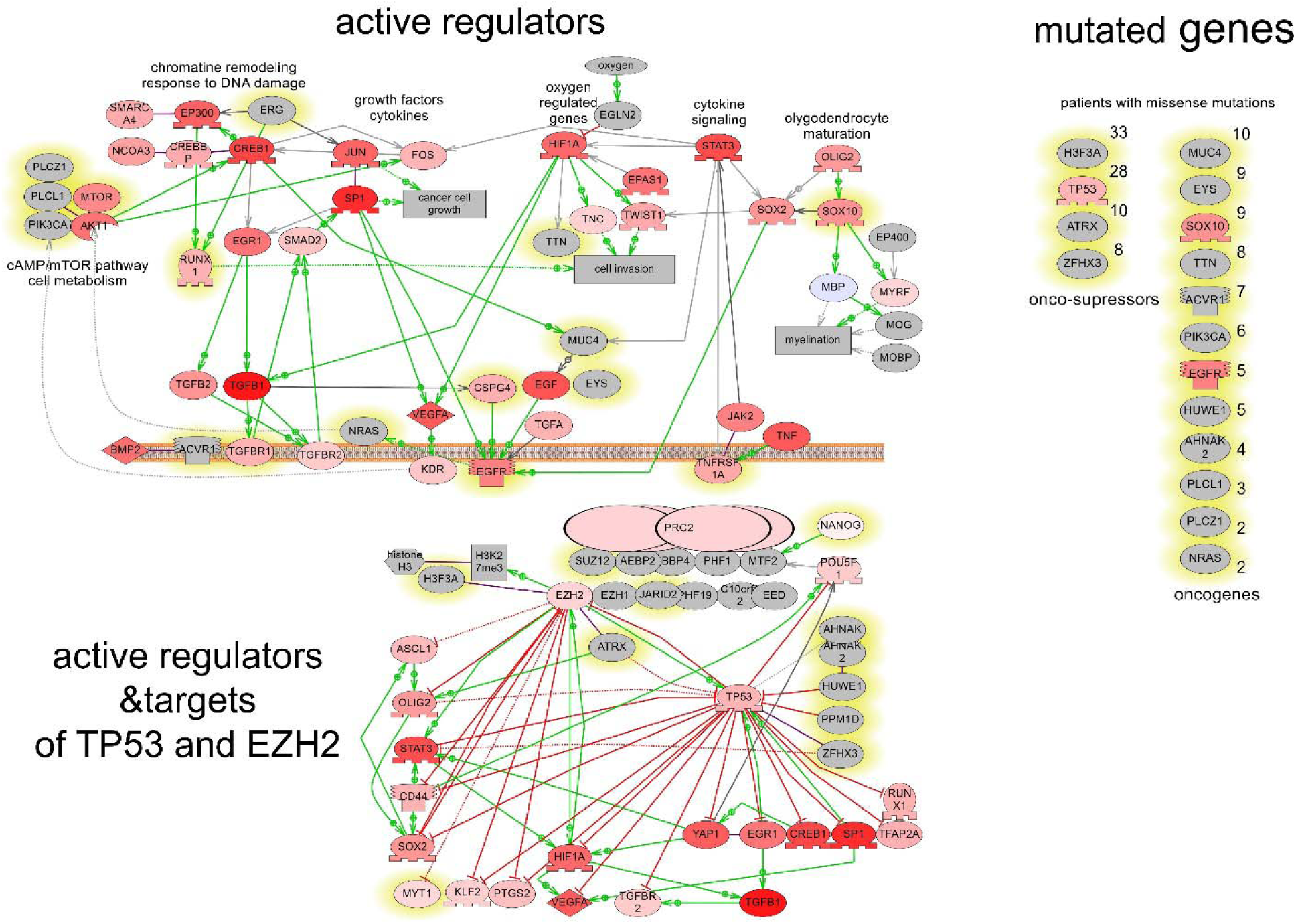
Predicted oncogenes in DIPG. Genes mutated in at least one patient are highlighted in yellow. Red color saturation shows activation score of active regulators.

Three proteins (RUNX1, SOX10 and EGFR) were both top scored active regulators and had mutations in exons in several patients (Figure B). SOX10 promotes expression of myelin and maturation of OPCs (García-León, et al., 2018). In glioblastoma model, SOX10 acted as tumor suppressor locking tumor cells in differentiated status (Brooks et al., 2021). Over-stimulation of EGFR is well known to cause multipotency and cell proliferation. RUNX1 can be oncogene or tumor suppressor depending on cancer type probably via interactions with WNT and SHH pathways (Li et al., 2021), it plays role in hematopoiesis and neuronal development and may promote glioblastoma cell growth (Sangpairoj et al., 2017).

There is an ambiguity in the use of terms “loss-of-function” and “gain-of-function” in the literature. In oncology “gain-of function” mutations promote cancer cell proliferation and migration, and “loss-of function” mutations repress cancer cell vitality. Same terms can be used in the literature to describe loss or gain of protein function. Mutations in TP53 gene mostly cause loss of its protein function (Carbonnier et al., 2020) but lead to “gain-of function” based on the oncology definition (Soussi et al., 2014) as wild-type TP53 has a protective role in cancer and is considered as onco-suppressor (discussed in the next section). Four of the genes mutated in DIPG patients (ANHAK1/2, HUWE1, PPM1D) may inhibit TP53 according to the literature and Elsevier Knowledge Network (Figure D). The clinical effects of such mutations in turn may depend on mutations in TP53, histone 3 and other oncogenes (discussed in the next section).

Additionally, together with TP53, mutations in H33A, ATRX, ZFHX3 genes also may play role of “gain-of function” mutations because these genes have protective role of onco-suppressors.

Top scored “regulators of suppressed sub-networks”, such as NCOR1, NROB2 and NEUROD1 can be considered tumor suppressors which are not active in DIPG tumor, because in healthy cells they are known to be involved in chromatin condensation during neuronal differentiation (Figure E). NEUROD1 is a master of cell differentiation, and its low activity may indicate strong pluripotency status of cancer cells in DIPG tumor. NEUROD1 was repressed by H3K27me3 in a mouse model of medulloblastoma and increasing NEIROD1 expression by pharmacological inhibition of H3K27me3/EZH2 reduced tumor growth (Cheng et al., 2020).

**Figure E.**
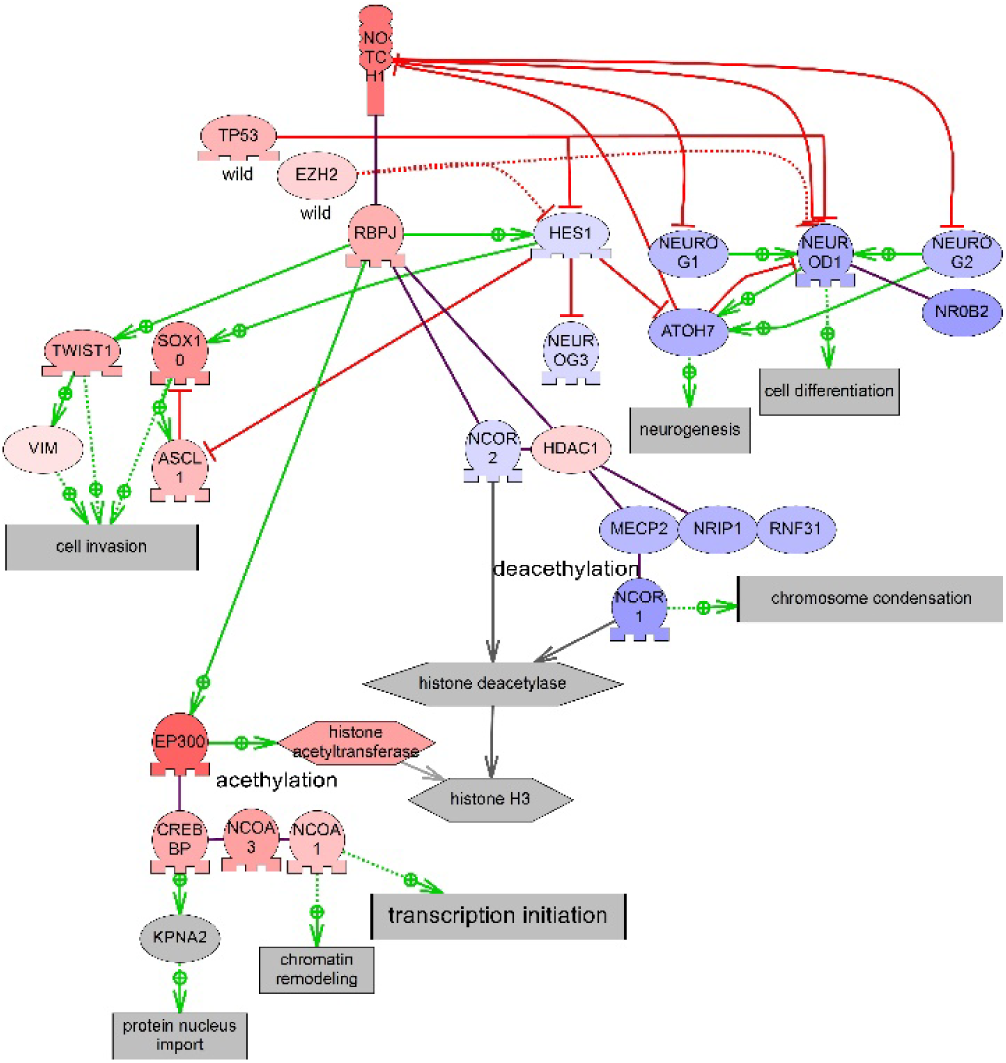
Suppression of NOTCH signaling in DIPG. Activity of regulators shown by red color saturation according to their activation score. Regulators of suppressed sub-networks are shown with blue color. The balance is shifted toward cell proliferation vs cell differentiation due to low activity of several NOTCH signaling expression targets.

In general, molecular cascades enriched with regulators of suppressed sub-networks in DIPG were linked to DNA acetylation and methylation, loss of neurotransmitter uptake, and to differentiation and myelination of neurons. Inhibited cancer hallmarks pathways were enriched with the top scored regulators of suppressed sub-networks regulators and down-expressed genes such as transcription factors from NOTCH pathway (Figure E), proteins in chromatin remodeling complexes (SCF/FBXW7, BAF) and DNA methylation complexes.

#### 1.3.4.2 Predicted changes in protein functions due to mutations in TP53 and histone 3 hypomethylation

H3K27M point mutation is considered a tumor driver in some DIPG types. While it is not entirely clear, why loss of trimethylation occurs in cells with H3K27M, it is hypothesized that this mutation increases the affinity of histone 3 to EZH2, a component of chromatin Polycomb repressive complexes 2 (PRC2), thus sequestering its methyltransferase activity (Brien et al., 2021; Harutyunyan et al., 2019; Lowe et al., 2019) and changing its ability to repress genes transcription through methylation (Cao et al., 2014; Justin et al., 2016).

In analyzed samples EZH2 and other two core subunits of PRC2 complex (EED, and SUZ12) were not a significantly differentially expressed gene neither in DIPG vs normal embryo cells nor in mutated TP53 vs wild TP53 comparisons (S. Figure 6). All subunits of PRC2 complex are essential for the methyltransferase activity of the complex, in addition to a histone chaperone RBBP7/RBBP4. However we focused on genes which are known to be regulated by EZH2 based on the previously reported assumptions that it is responsible for early phases of oligodendrocyte differentiation as a consequence of the H3K27M mutation. (Wang et al., 2020).

Using Elsevier Knowledge Graph (EKG) and results of transcriptomics analysis, we identified 140 differentially expressed genes that could be over-expressed in DIPG samples because of their hypomethylated status which is linked to lack of PRC2/EZH2 activity (Laugesen et al., 2019). To find them we considered reported in the literature and present in the EKG database interactions with suppressing effects between EZH2 and other genes. As EZH2 targets, only genes with major differences in expression were considered (Supplemental file “EZH2 predicted expression targets”). DNMT3A, CCND1, RUNX3 were among EZH2 expression targets supported with the greatest number of references in the network. 59 EZH2-dependent regulators of gene expression included markers for OPCs cells, and regulators with highest scores (TGFB1, STAT3, CTNNB1 and others).

PCR2 forms multiprotein complexes with many chromatins remodeling complexes, methyltransferases and histone acetylases to regulate gene expression. For example, for DIPG it was shown that EZH2 is regulated by phosphorylation at Ser21, Thr311, and Thr487 by signaling protein kinases (AKT, AMPK, and CDK1, respectively) causing dissociation of PRC2 subunits and the reduction of H3K27 methylation (Duchatel, 2019). Also, it was reported that H3K27M-glioma overexpressed BMI1, the subunit of another Polycomb complex PRC1 and it was sensitive to its inhibition. PRC1 mono-ubiquitylates lysine 119 on histone H2A (H2AK119ub1) and there are many cross-interactions between PRC1 and PRC2 (Fiblin, 2018). In addition, BMI1 inhibited PRC2-mediated methylation of histone H3 in vitro (Cao et al., 2014). In our analysis BMI1 had the highest average level of expression (DE 13.1 to 10.8 in all samples, in interval [13.3; -13.8]) in DIPG samples but was not a significant regulator. We suggest that the activation of PRC1 complex may occur because of the feedback compensatory mechanism for PRC2 dysfunction.

PRC2 complex needs additional regulators targeting PRC2 to chromatin and stabilizing the complex. One of them, MTF2 (PCL2) can mediate de novo recruitment of PRC2 by binding to unmethylated CpG islands in condition of unbound DNA (Healy, et al., 2019; Mierlo et al., 2019). We found that MTF2 and C10orf12 were down expressed (DE 0 to -8.3 and 0.5 to -2.4 in all samples, in interval [13.3; -13.8]) in DIPG tumors vs control as well as their transcription factors (NANOG and POU5F1) (Supplemental file “PNOC003vsGSE120046”). Jumonji family histone demethylases are also involved in maintenance of hypomethylation of H3K27 and other genes in DIPG (Franci et al., 2014; Nikolaev et al., 2020). Jumonji family histone demethylases KDM6A (UTX) and KDM6B had high level of average DEs and were in the list of regulators in analyzed samples.

We combined the list of EZH2-dependent targets with genes that may have transcriptional changes because of mutations in TP53. First, we suggested that transcription targets whose expression patterns in DIPG samples were not consistent with effects of wild-type TP53 reported in the literature may indicate the effects of mutant TP53. We found 81 probable transcriptional targets of wild-type TP53, 32 of them were present in the literature network as targets activated by TP53, however they were down-expressed in DIPG samples. The rest 49 were reported to be inhibited by TP53 but were over-expressed in analyzed samples. 9 targets were reported to control OPCs development and 19 were regulators of gene expression in DIPG patients (Supplemental file “TP53 predicted expression targets”). Connections with BCL2, MYC, SP1 are supported by the greatest number of references (96, 49 and 45 accordingly).

Only 13 predicted expression targets for TP53 overlapped with EZH2/PRC2 predicted targets (S. Figure 7). These were SOX2, CD44, KLF2, PTGS2, HIF1A (active regulators), DNTM3A, SP7, DICER1 (significant nonactive regulators), BCL2, MYC, CHUCK and CDH1 (not significant regulators).

We checked if the patterns of the expression of TP53 and EZH2-depended targets had differences in samples with mutant TP53 versus DIPG samples with no mutations in TP53 (S. Figure 8). We found 27 genes, targets of TP53 and EZH2 with different levels of expression in samples with mutations in TP53 (all samples had mutations in histone 3).

All findings were combined into the model depicted on Figure F titled “Mutated TP53 effects on gene expression in DIPG”. Levels of nine targets known to be suppressed by wild TP53 (HIF1A, MYC, TGFBR2, NOS2, SCL2A1, SLC7A11, AR, CDKN2A) were in general higher in samples with TP53 mutations (see also S. Figure 8). So, they could be also targets of mutant TP53 in DIPG tumor acting as oncogenes because they can promote cancer growth directly or through maintaining angiogenesis or lack of T-cell immunity. On the contrary, expressions of three targets known to be activated by wild TP53 targets (TFAP2A, GAD2, NEAT1) were lower in samples with TP53 mutations. At least one of them, TFAP2A can be a probable onco-suppressor which activity attenuated due to mutations in TP53. Some likely changes in cell processes linked to mutations in TP53 and reconstructed models are discussed below.

**Figure F.**
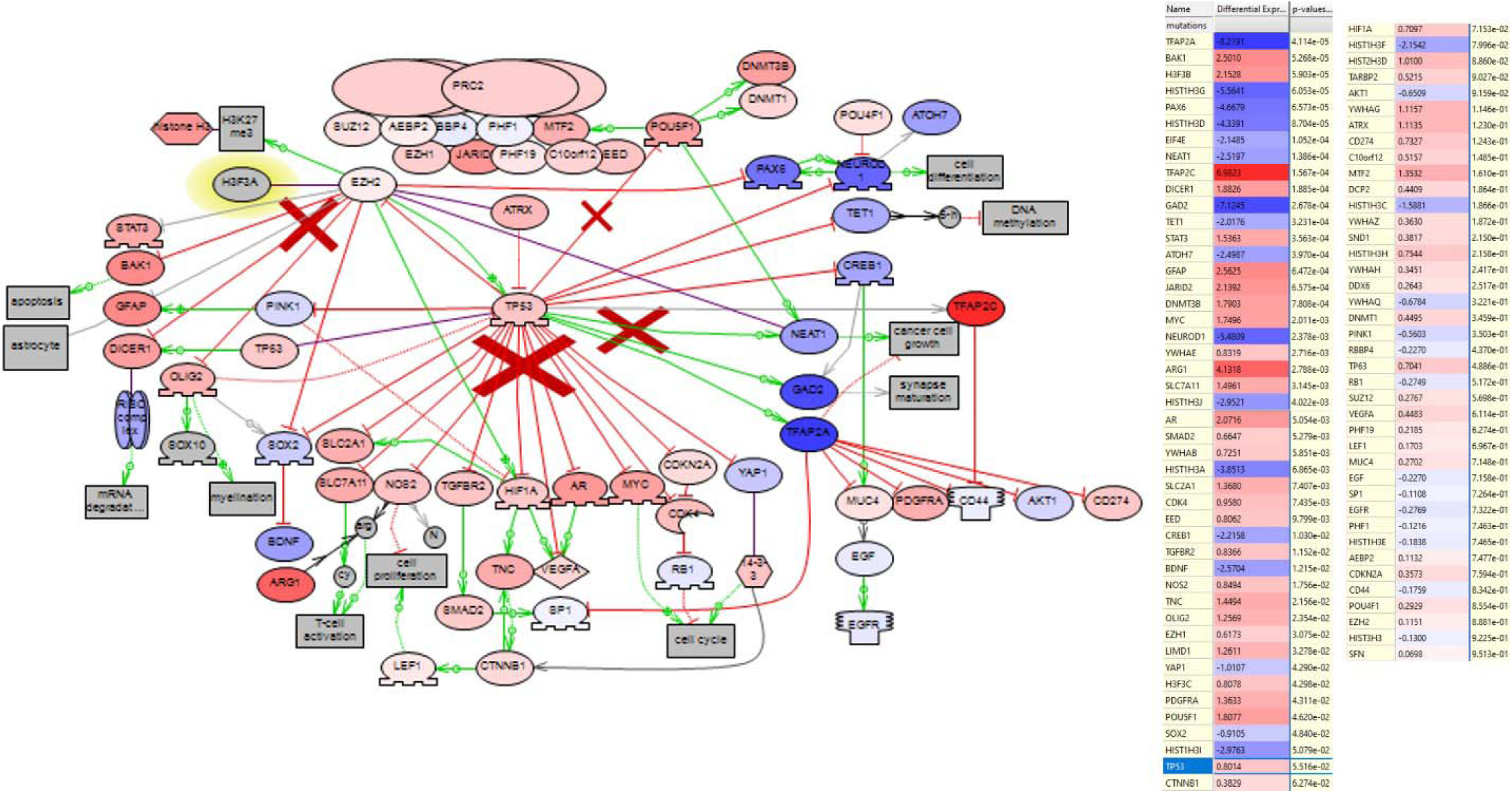
Differential expression of TP53 and EZH2 targets in DIPG samples with mutant TP53 versus samples with no mutant TP53 is shown by color saturation. Tables on the right show logarithm of fold changes of gene differential expression between DIPG samples and normal human pons sorted by p-values. The color of arrows between genes indicates regulation effect of protein-protein interaction most often discussed in literature. Red arrows indicate negative effect (one protein inhibits another), green – positive effects. HIF1, SOX2, CD44, GFAP, YAP1, CREB1, TFAP2A, and POU5F1 were found to be active regulators of over-expressed genes in DIPG vs embryonal cells based on sub- network enrichment analysis. NEUROD1, DICER1, GAD2 and TFAP2C are regulators of suppressed sub-networks. EZH2 is regarded as a suppressor of gene expression due to its ability to methylate gene promoters. However emerging literature suggests that EZH2 can also directly activate transcription of some genes such as STAT3 (Tabbal et al., 2019).

##### Low response to oxygen stress

Hypoxia inducible transcription factor HIF1A was one of the top scored active regulators that was mainly overexpressed in DIPG samples with mutant TP53 and suppressed by TP53 in normal cells according to literature network. EZH2 was shown to be an activator of HIF1A expression (due to its suppression of EAF2 expression) (Pang et al., 2016). HIF1A is a regulator of low oxygen stress response including activation of misfolded protein and mitochondrial response. HIF1A-related cascade may be a reason for tumors radio resistance, since oxygen is recognized as important factor in the cytotoxic effects of radiation (Harada, 2016). Recently, mutant TP53 was reported to cause radio resistance in H3-K27M–mutated DIPG (Werbrouck et al., 2019). Other proteins related to oxygen or proteolysis cascades in neuronal tissues were also highly overexpressed in DIPG samples. Among them mitochondrial PINK1 which is inhibits HIF1A in normal cell and itself is a negative target of TP53. Animal models showed that PINK1 is important for astrocyte differentiation and proliferation probably through activation of EGFR/AKT cascade or GFAP expression (Barodia et al., 2019; Choi et al., 2016). Finally, in addition to hypoxia cascade, wild-type TP53 suppresses VEGFA expression directly and via inhibition of HIF1A (Farhang Ghahremani et al., 2013). In DIPG samples PINK1 and VEGFA, as well as other genes associated angiogenesis cascades, such as NOS2, were over-expressed compared to control (supplemental materials, file 2).

##### OPCs pluripotency biomarkers

Changes in OPCs expression markers should be directly involved in DIPG mechanisms. Overexpression of SOX2 and Nestin may indicate pluripotent status of OPCs or undifferentiated neural progenitor cells (Pajovic, et al., 2020; Robins et al., 2013). Transcription factor SOX2 was indeed overexpressed in most DIPG tumors compared to control (4.8; 1.2 DE log-changes in interval [13.3; -13.8]). SOX2 was not suppressed both in samples with mutated TP53 and in samples without TP53 mutations (Figure F, S. Figure 8), which suggests that SOX2 expression is not dependent on TP53 mutations. Our calculations show that SOX2/SOX10/OLIG1/OLIG2 signaling is hyperactivated in the analyzed samples. We also found that SOX2, Nestin, OLIG1/2, GFAP and ASCL1 can be downstream targets of H3K27me3 and TP53 (Figure F) according to the literature. In conclusion, SOX2 together with SOX10 and OLIG2 were top scored predicted active regulators of gene expression in DIPG tumors and may regulate EGFR signaling during the myelinization and maturation of OPCs. SOX10 may play more important role in DIPG since it was mutated in 9 patients.

##### Cell cycle

Wild type TP53 inhibits MYC expression to induce cell cycle arrest (Wagner, et al., 1994). In human medulloblastoma, MYC was identified as a major downstream target of EZH2 and shown to be essential for stemness and tumorigenicity (Suva et al., 2009). MYC amplification initiated medulloblastoma formation in combination with TP53 loss and overexpression of chromatin remodeling protein REST/NRSF (Swartling et al., 2014). In analyzed DIPG samples, MYC has higher level of expression compared to control, however, it was not a significant regulator of other genes.

Expression of other proteins involved in cell cycle regulation also may depend on TP53. Recently YAP and mutant TP53 were shown to have common transcriptional program for targets primarily involved in cell cycle regulation. Moreover, different TP53 GOF mutants (p53R280K, p53R175H, p53A193T, p53R248L, p53R273H, p53L194F, and p53P309S), but not wild-type TP53 protein, can physically interact with YAP (Di Agostino et al., 2016). YAP was over-expressed in DIPG samples in comparison with control, but its expression probably does not depend on mutations in TP53 (S. Figure 8). Also PRC2 complex was reported to repress TP53 and its target gene p16INK4a (CDKN2A) in OPCs (Wang et al., 2020), however in mice models it was shown that CDKN2A expression was independent of EZH2 hypomethylation of H3K27 (Cordero et al., 2017). In analyzed DIPGs samples, CDKN2A and other controlled by TP53 inhibitors of cell cycle were slightly overexpressed compared to normal tissues or to samples with TP53 mutations.

##### Other TP53 targets

Wild TP53 acts as tumor suppressor not only by inhibiting expression of oncogenes but also by inducing expression of tumor suppressors such as TFAP2A. TFAP2A is involved in the inhibition of cell cycle and cancer growth. Both upregulation and downregulation of AP2 transcription factors (TFAP2C and TFAP2A) has been observed in multiple human cancers (Kołat et al., 2019). AP2 transcription factors may be affected by mutant TP53 in DIPG samples, since they appear to have tendency to change their expression in samples with mutant TP53 vs samples with wild TP53 (S. Figure 8). TFAP2A signaling also linked to CD44 hyaluronan receptor which has a prominent role in the maintaining cancer cell growth. CD44 expression probably does not depend on mutations in TP53, however it is overexpressed, and it is an active regulator of gene expression in DIPG in our analysis (Figures D, F). The pathway “Hyaluronic Acid, CD44 and HMMR in Cancer Cells Motility, Invasion, Proliferation and Survival” summarises effects of CD44 signalling which includes many active regulators from previous analysis (SOX2, POU5F1, STAT3, EGFR) (S. Figure 9). In addition, we found the impairment of metabolic regulation pathways in DIPG. Correlation analysis of DE genes revealed several clusters where central top over- expressed proteins were tet methylcytosine dioxygenase 3 (TET3) and ubiquitin ligase CUL4B. TET3 was important for the regulation of H3K27me3 in several genes in cancer cells (Cao et al., 2019). CUL4B is known to be activated by ER and oxygen stress. It degrades TP53 via proteosomes and also promotes H2AK119 monoubiquitination and trimethylation of H3K27 in embryonal cells (Hu et al., 2012; Wei et al., 2015). We propose that the overexpression of TET3 an CUL4B in analyzed samples in comparison to embryonal cells may indicate the presence of the feedback loop in response to mutant TP53 accumulation. Altered expression of metabolism-related genes is well studied in cancer and is targeted by drugs explored for DIPG (Lin et al., 2019). We could not find differences between samples with mutated H3.1 and samples with mutated H3.3A. We could not confirm that p38 MAPK, PI3K, or Rho GTPase pathways were central to H3.1K27M related DIPG pathobiology as was reported earlier (Nagaraja et al., 2019).

To further support our model, we compared expression of genes and regulators in different patients. Samples related to patient P34 (deletion in TP53), P02 (hotspot missense SNV in TP53, rs28934575), P21 (hotspot missense, rs121913343) were similar to each other and to average scores distribution, while P12 (with rs483352697) has lowest similarity to average scores distribution. In P34 sample with deletion in TP53 and mutations in H3.3A we found 9 predicted active regulators. Sample P01 with wild TP53, had only 2 active regulators (Figure G). This observation supports the model “Mutated TP53 effects on gene expression in DIPG” and our hypothesis that tumor growth in P34 may depend on mutant TP53 and hypomethylation due to lack of EZH2 activity.

**Figure G.**
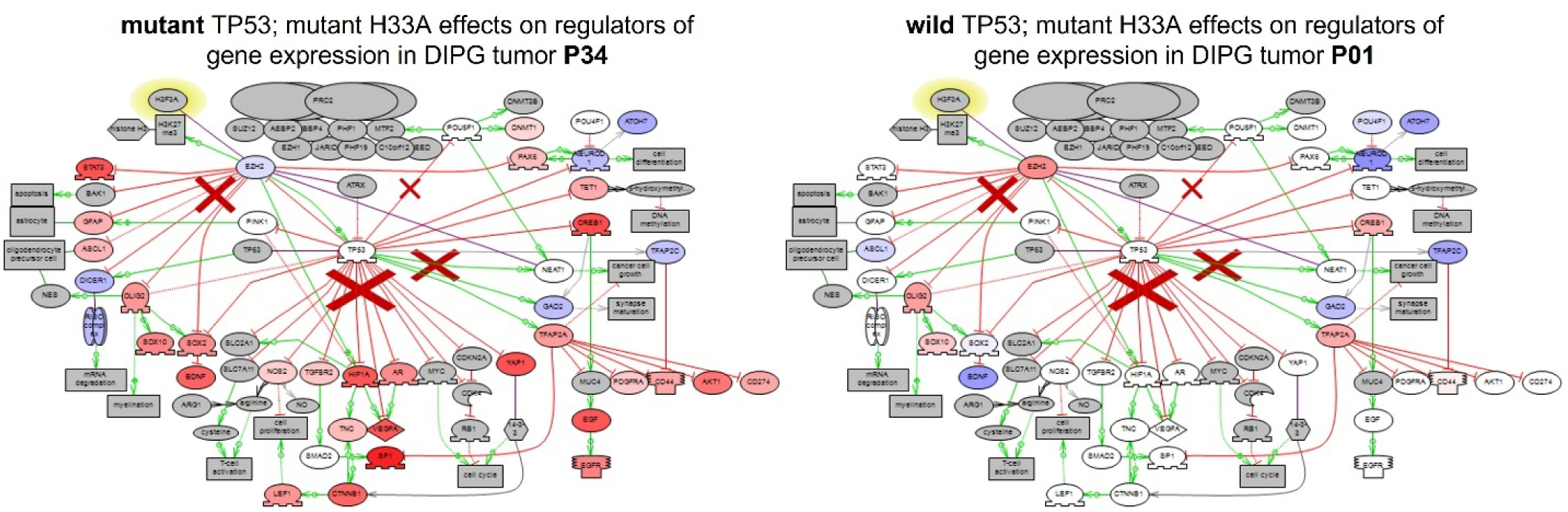
Predicted role of mutations in TP53 and H33A in molecular mechanisms of the DIPG tumor growth. Color of entities shows the average regulators activity scores in 21 DIPG samples with mutated TP53. P034 sample has deletion in TP53 and mutations in H33A. P01 sample only has mutations in H33A. Active regulators are highlighted in red; regulators of suppressed sub-networks - in blue; not significant - in grey. Arrows between genes/proteins show protein-protein interaction most often discussed in literature. Red arrows show inhibition, green – activation.

To summarize, we used results of “pathway activity analysis” of 33 DIPG tumors to find oncogenes and onco- suppressors that may be driven by H3K27me3 and TP53 mutations in OPCs. We suggest that several major transcription factors, including HIF1A, POUF51, SOX2, OLIG2, may be under control of H3K27me3 and mutated TP53, and may be possible key regulators of DE genes in DIPG (Table 2, Supplemental file 11). These genes regulate oligodendrocyte fate, cancer cell metabolism and cancer vascularization.

**Table 2.**
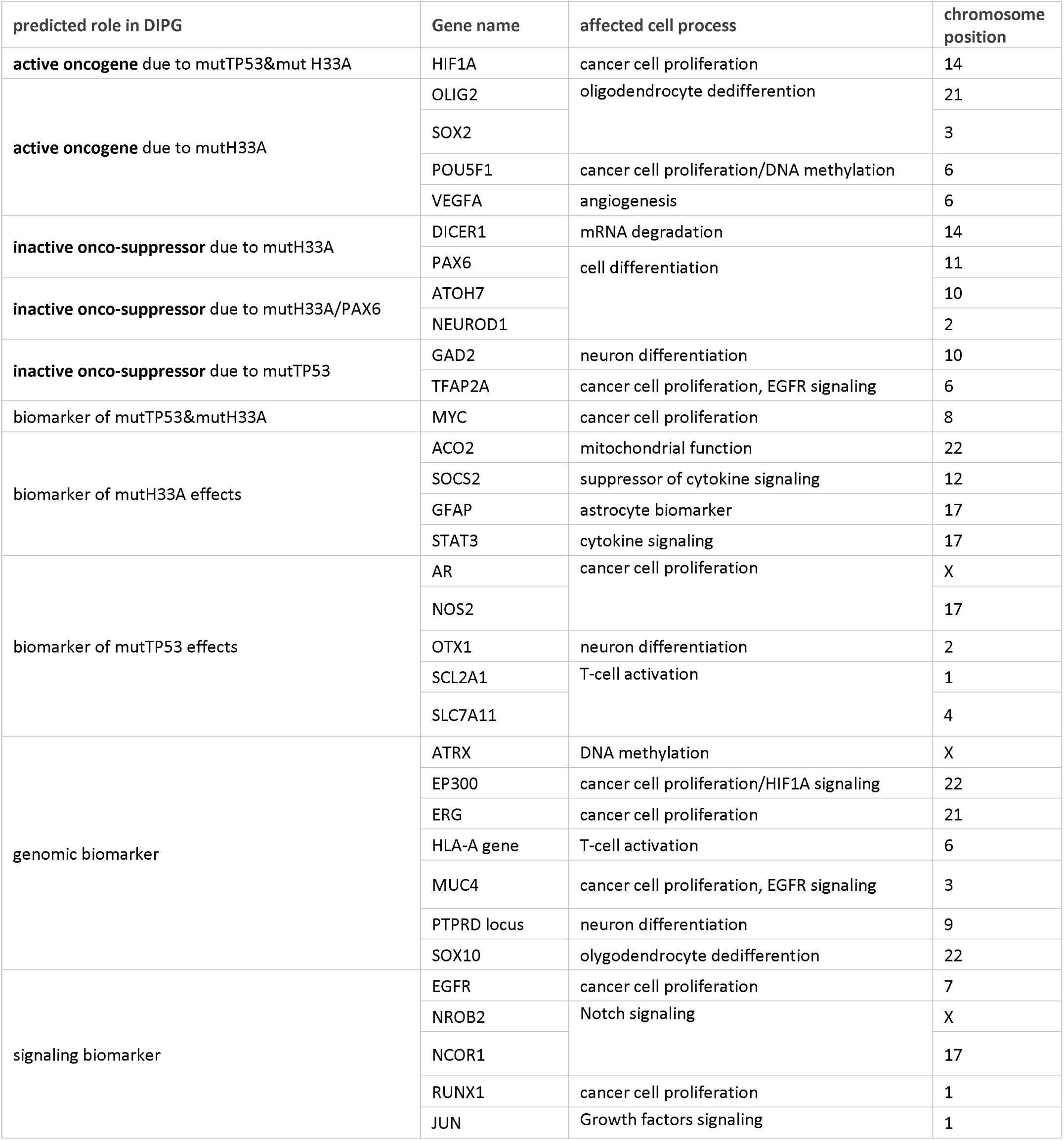
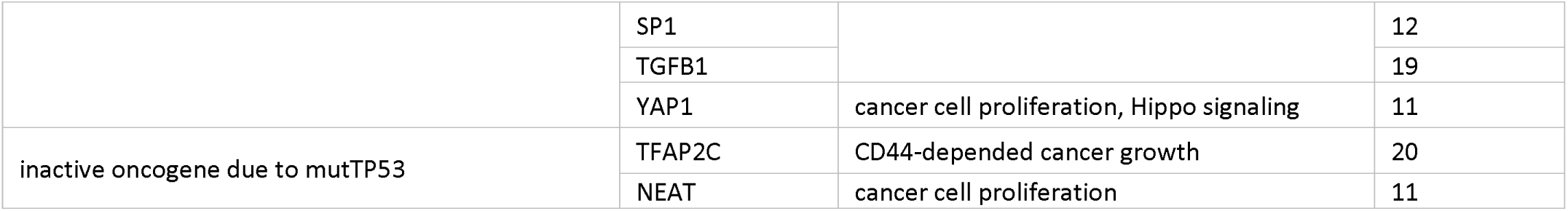
Predicted regulators and biomarkers of DIPG subtype with histone 3 and TP53 mutations.

### 1.3.5 Repositioning of FDA approved drugs for DIPG treatment

Development of DIPG treatments is an area of active research (Lin, et al., 2019). We found 169 drugs in the Elsevier Knowledge Graph that were tested in clinical trials for DIPG treatment. 61 clinical trials with drugs for DIPG are registered at clinicaltrials.org, and 20 drugs have completed Phases 1 to 3 trials (S. Figure 10). Among them, panobistat, palmociclib, temozolomide, bevacizumab, nimotuzumab were more often discussed in published results.

We used SNEA to find drugs which may inhibit major regulators of aberrant gene transcription in DIPG and TP53- dependent regulators (see Methods). Drugs were searched among a collection of 22,150 substances, including FDA-approved drugs and investigational compounds. Drugs were scored by SNEA/GSEA enrichment score and by the number of inhibited regulators (Supplemental file “Drugs by SNEA”). Proteosome inhibitor MG132, DNA methylation inhibitor zebularine, and PI3K inhibitors (LY294002, wortmannin) had the highest scores because they were reported to inhibit all 117 DIPG regulators. Other top scored drugs and drug candidates used for treatment of breast cancer and compounds known as inhibitors of MAPK cascade, EZH2, DNA methylation and HDACs. The list included many natural compounds studied as anti-cancer, anti-inflammatory, or neuroprotective agents. From total number of 143 compounds predicted by SNEA, 41% of compounds have been studied as anti- cancer treatments, and 13% were FDA approved anti-cancer drugs (Figure H).

**Figure H.**
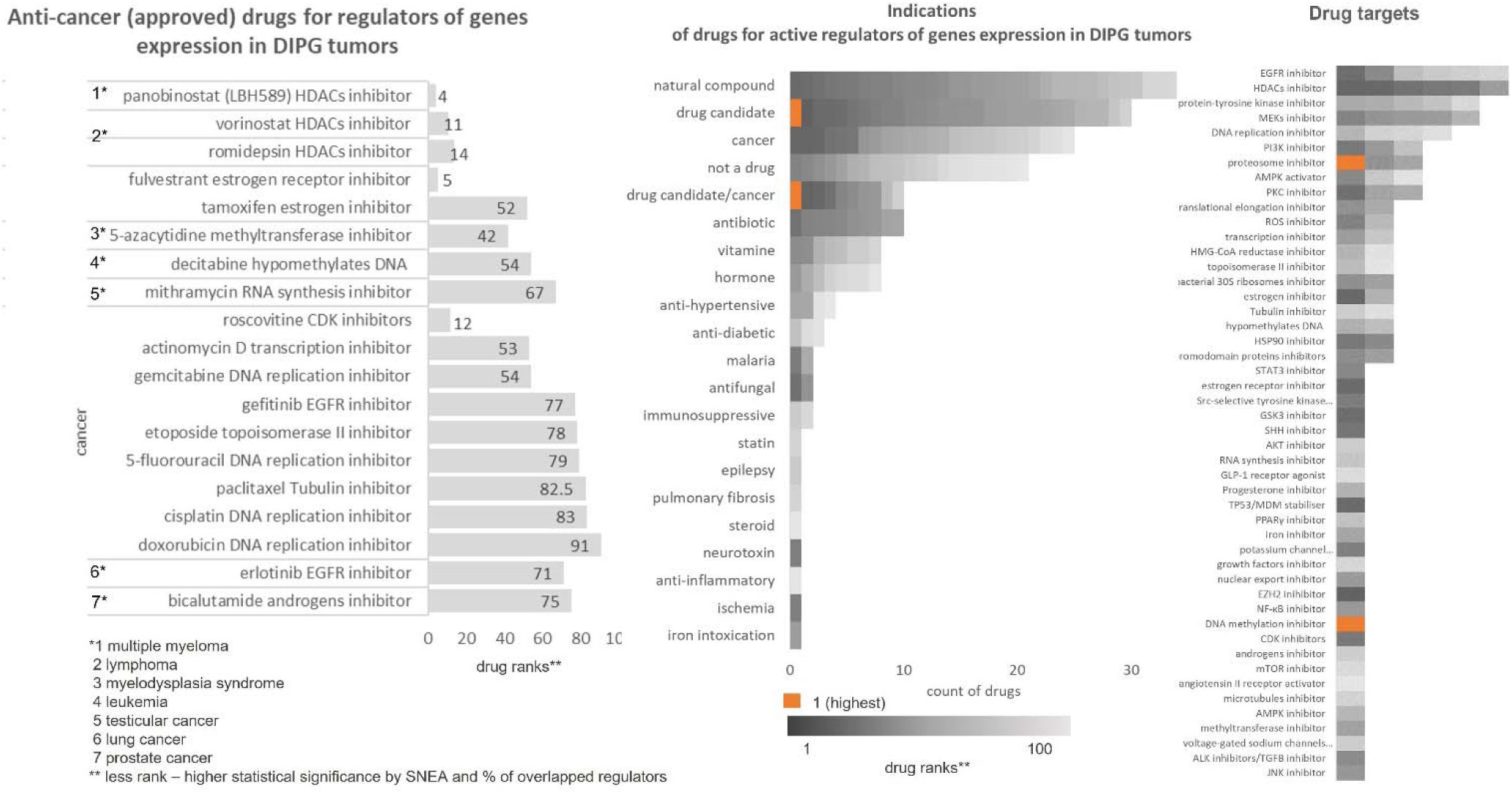
Anti-cancer drugs found by SNEA for active regulators of gene expression in DIPG.

Panobinostat (LBH589) appeared to be to score anti-cancer drug based on our analysis. Panobinostat is one of the most clinically tested drugs (Long W, 2017). It is a multi-HDAC inhibitor which inhibited DIPG cell lines in an in-vitro screen and repressed global histone h3K27Ac acetylation caused by H3K27M and thus may enhance activity of PCR2 complex (Brien GL, 2021).

Since every patient has an individual pattern of mutations and key regulators, it is reasonable to search drugs capable of reversing disease transcriptome state individually for each patient. We used SNEA to find potential drugs for patient P034 with deletion in TP53. The list included 23 compounds with anti-cancer activity. Top scored drugs were LY294002 (drug candidate, PI3K inhibitor), U0126 (anti-cancer drug candidate, MEKs inhibitor), antiproliferative metabolic drug metformin 3-deazaneplanocin A, losartan (Angiotensin II receptor inhibitor), actinomycin D (anti-cancer), imatinib (anti-cancer) (S. Figure 11). The complete list of drugs that can be repurposed for this patient is provided in Supplemental file “Drugs by SNEA”.

Finally, focusing on the TP53 role in DIPG, we found drugs reported in literature capable of inhibiting TP53 with mutations. We trained Elsevier Natural Language Processing technology to recognize sentences reporting drugs inhibiting “mutant TP53” and applied it against standard literature corpus including all PubMed abstracts, all full-text articles published by Elsevier biomedical journals or available from PubMed Central. This effort yielded 585 compounds reported to inhibit “mutant TP53” (Supplemental file “Drugs for mutant TP53”). 28 drugs had been reported to interact with TP53 directly. Among them an anticancer drug Doxorubicin (Adriamycin) and arsenic compounds (for example Trisenox, which are toxic natural compounds that may be used for cancer treatment) had the highest number of references. Mechanism of direct inhibiting of TP53 with mutations should be specific to the nature of the mutation. That is why we focused on drugs that less depends on type of mutations and can level out effects of non-functional TP53 on downstream cell signaling indirectly. We filtered compounds reported in the literature to be able to inhibit “mutant TP53” by their ability to cross blood-brain barrier which we defined by presence of interactions in Elsevier Knowledge Graph between a compound and neurological side-effect (terms “neurotoxicity”; “central nervous system toxicity”; “delayed neurotoxicity”’ “neurotoxic disorder”; “neuromyotoxicity”; “neurosensory toxicity”).

Next, we filtered drugs by ability to inhibit heat-shock response necessary for mutant TP53 stabilization. The following cell processes were considered relevant:

(a) inhibitors of components of cytosolic chaperone complex: HSP90 (e.g., Geldanamycin, Tanespimycin) and HSP40 (e.g., Statins). We found 8 different statins (atorvastatin, simvastatin, lovastatin, etc.) in the list. Statins have been shown to inhibit mutant TP53 by down-regulating biosynthesis of phosphomevalonate, which is necessary for mutant TP53 stabilization by DNAJ/HSP40 protein (Tutuska et al., 2020). It was previously demonstrated that statins delocalized YAP, probable TP53 expression target by our study, in the cytoplasm thereby severely impairing its oncogenic activity (Sorrentino et al., 2014).

(b) activators of chaperon-induced autophagy (e.g. Tozasertib, Myfortic) which leads to increased degradation of mutant TP53 in cytosol and subsequent decrease of its nuclear concentration (Vakifahmetoglu-Norberg, et al., 2013).

(c) inhibitors of mortalin (HSPA9) (e.g., Embelin, Withaferine A). Mortalin is a chaperon protein which targets both wild-type and mutant TP53 proteins for degradation. Presumably, inhibition of mortalin increases wild type to mutant TP53 ratio, because wild type protein is more stable (Sundar et al., 2019).

(d) inhibitors of heat-shock transcription factor HSF1 (e.g., Quercetin, Imatinib). HSF1 is activated by mutant TP53-induced heat-shock response to increase production of HSP90-HSP70-HSP40 cytosolic complex stabilizing mutant TP53 (Boysen, et al., 2019).

(e) HDAC inhibitors (e.g., Panobinostat, Romidepsin). HDAC1/2 are directly activated by HSF1 which may lead to de-repression of PRC2 complex inhibited by H3K27M mutation. HDAC6 also forms a repressive complex with HSP90 and inhibits HSF1 transcriptional activity thus controlling the duration of heat-shock response (Pernet et al., 2014). HDAC inhibitors can cause HDAC6 dissociation from inhibitory HDAC6/HSP90/HSF1 to activate HSF1 and heat-shock response that stabilizes mutant TP53. However, in cytoplasm pharmacological inhibition of HDAC6 can destabilize mutant TP53 (Muller and Vousden, 2014).

We ranked filtered out drugs that reported to be able inhibit “mutant TP53” indirectly (262 drugs in total) by number of references supporting the relations between a drug and “mutant TP53” and clustered them by various cancer-related indications (Supplemental file “Drugs for mutant TP53”, Table 2). Drugs with the highest scores were anti-cancer HDACs inhibitors, DNA damaging factors, and tubulin inhibitors (Supplemental file “Drugs for mutant TP53”, Table 2). Vorinostat, cisplatin and paclitaxel were top ranked drugs capable of inhibiting mutant TP53 via HSP90, HDACs, SIRT1 or HSF1.

**Table 2.** Drugs with predicted effects against mutated TP53.

| Mechanism of action | Drug | Normalised score (1-100) | Number of references |
| --- | --- | --- | --- |
| **HDACs inhibitor** | **Sum** | **168** | **3494** |
|  | vorinostat | 100 | 2110 |
|  | trichostatin A | 42 | 870 |
|  | HDAC inhibitors | 14 | 272 |
|  | MS-275 | 12 | 242 |
| **DNA damage response (DDR) inhibitors** | **Sum** | **162** | **3387** |
|  | cisplatin | 63 | 1322 |
|  | cyclophosphamide | 50 | 1047 |
|  | gemcitabine | 49 | 1018 |
| **tubulin inhibitor** | **Sum** | **104** | **2178** |
|  | paclitaxel | 61 | 1282 |
|  | docetaxel | 43 | 896 |
| **topoisomerase II inhibitor** |  | **70** | **1450** |
|  | doxorubicin | 44 | 908 |
|  | etoposide | 26 | 542 |
| **EGFR inhibitor** | **Sum** | **52** | **1069** |
|  | erlotinib | 35 | 716 |
|  | gefitinib | 18 | 353 |
| **VEGFA inhibitor** | **Sum** | **52** | **1079** |
|  | bevacizumab | 52 | 1079 |
| **mTORs inhibitor** | **Sum** | **50** | **1029** |
|  | everolimus | 31 | 630 |
|  | rapamycin | 20 | 399 |

## 1.4 Limitations

Our findings have some limitations. The number of patients is relatively small. Embryonal cells were used as controls to calculate DE, not healthy brain tissue from the same person or a person of matching age. Data in the Knowledge Graph used for the analysis includes information from animal studies which can differs from human cell and cancer biology. Pathway activity analysis tends to bias towards most studied proteins and genes (proteins with higher connectivity in the database). For example, novel TGFB inhibitors, such as vactosertib or fresolimumab, were not found by our approach due to a relatively small number of publications about their mechanism of action which translates into low connectivity in the Knowledge Graph.

## 1.5 Methods

### 1.5.1 Data from OpenPBTA

We used RNAseq data from 32 samples obtained from 30 DIPG patients in OpenPBTA (https://www.ccdatalab.org/openpbta). Analysis of type and location of mutations was performed using Python, UNIPROT and PFAM public databases (https://github.com/pbnjay/lollipops/blob/master/README.md).

To calculate differential expression between DIPG tumor and normal tissue, we used data of 17 OpenPBTA DIPG patients with mutation in histone 3 that also have mutations in TP53 gene (P02, P05-P14, P21, P27-P28, P30- P31, P34-P36). Patients P01, P06, P017, P019, P023, P026, P033, P038 did not have mutations in TP53 and were excluded from calculation of average DEs. Two more patients with H3K27M mutation in H3.1 gene and with mutation in TP53 gene (P16, P20) were also added for average DE calculation. We also have included in analysis 3 patients without mutations in H3.3A gene but with mutations in TP53 (P4, P11, P37). Samples were collected twice for two of the patients (P06 and P07), before and after radiation therapy because they did not respond to the therapy and their disease had progressed.

### 1.5.2 Differential expression

To obtain the best control for calculation of differential expression, we considered three public datasets measuring normal brain tissue: single-cell transcriptome dataset of embryonic pons (GSE120046) (Fan et al., n.d.); 16 weeks embryonic brain pons from Human Developmental Biology Resource (HDBR) (https://www.ebi.ac.uk/arrayexpress/experiments/E-MTAB-4840) (Lindsay et al., 2016); and postmortem brain cerebellum data from GTEx consortium (https://gtexportal.org/).

We calculated transcripts per million for every gene and compared differential expressions between each DIPG patient’s sample and normal brain tissues with the DESeq2 package (Love, et al., 2014). Then we tested three versions of differential expression with cancer hallmarks pathways enrichment scores. GSE120046 dataset measuring single cell transcriptome of embryonic pons produced the smallest GSEA p-values for all active cancer hallmarks in several patients and therefore was selected for analysis of all patients. GSE120046 transcriptome datasets of embryonic pons from combined data from 4278 individual pons cells from embryos ranging from gestational week 9 to 28. According to the authors their sample included neural progenitor cells, cortical excitatory neurons, excitatory neuron-like cells in the pons (pons neu), Cajal-Retzius cells, cortical interneurons, astrocytes, oligodendrocyte lineage cells, immune cells (microglias, macrophages, and T cells), and endothelial cells and blood cells. We used all types of cells from pons since DIPG tumor samples also contain the mixture of various cell types.

### 1.5.3 Sub-Network Enrichment Analysis (SNEA)

Sub network enrichment analysis (SNEA) is the algorithm in Pathway Studio (Nesterova, et al., 2020; Sivachenko et al., 2007). Similar algorithms are also called causal reasoning in the similar software package (Krämer et al., 2014). In this work we have used SNEA for find statistically significant sub-networks with protein regulators upstream of differentially expressed genes and to find statistically significant sub-networks with drugs upstream of proteins. SNEA uses expression relationships between genes in the knowledge graph to find upstream regulators of differentially expressed (DE) genes. The activation score of a regulator is calculated as difference between concordant and discordant changes in differential expression of its biomarkers normalized by square root of total number of regulator targets (https://service.elsevier.com/app/answers/detail/a_id/4530/supporthub/pathway-webmammal/kw/activation). Concordant changes are consistent with the positive or negative effect of a regulatory relationship between target gene and regulator gene.

Since the data model in Pathway Studio has more than 172 types of semantic relationships SNEA allows selecting different sub-network configurations to answer various biological questions (https://service.elsevier.com/app/answers/detail/a_id/3014/supporthub/pathway-webmammal/kw/relations). Different sub-network configurations can be further filtered by built-in Pathway Studio Ontology to select only sub-networks formed by certain categories of entities, e.g., only drugs or only transcription factors. To predict regulators of genes expression we used knowledge graph relationships “Expression” and “PromoterBinding”. To predicted drugs with SNEA we used the activation scores of regulators of expression as input and SNEA was configured to find only drugs inhibiting its protein targets (all possible types of relationships). The more regulators are inhibited by a drug and the higher the average activation score of its targets the higher drug potential to inhibit differential expression in the tumor. Negative “activation score” indicates that drug inhibits active regulators; positive “activation score” indicates that drug inhibits regulators of suppressed sub-networks.

### 1.5.4 Gene-Set Enrichment Analysis (GSEA)

GSEA algorithm in Pathway Studio allows to identify user-defined gene sets, ontology groups and pathway collections statistically associated with an input list of genes. We used Elsevier Pathway Collection and list of genes with differential expression or list of predicted regulators of expression as an input for GSEA (Subramanian et al., 2005). Pathways are considered as gene sets by GSEA and are ranked by p-value.

### 1.5.5 Knowledge graph analytics and model construction

Data was analyzed using Pathway Studio (www.pathwaystudio.com) software from Elsevier, which allows access to Elsevier Biology Knowledge Graph extracted by Elsevier’s proprietary natural language processing (NLP) technology. Elsevier processes with its NLP entire PubMed (>26 mln abstracts) and more than 5 mln full-text biomedical articles to generate the knowledge graph containing more than 12 mln unique relations between biomedical concepts. Elsevier’s knowledge graph covers interactions between different concepts, such as biomolecular and drug-target physical interaction networks, the regulatory network between genes, proteins and substances, and high-level abstract concepts such as diseases, cell processes, cell types and clinical parameters that are used by scientists to describe biological processes in scientific literature.

DIPG models were manually designed using Pathway Studio software to depict probable disease mechanisms based on relationships covered by Elsevier Biology Knowledge Graph or added manually based on literature review. The method of reconstruction of disease models is described in (Nesterova, 2020).

### 1.5.6 Supplemental materials

Supplemental materials can be downloaded in ResearchGate (DOI: 10.13140/RG.2.2.36479.07848).

Pathways from Elsevier Pathway Collection can be viewed by links at Bioportal (https://bioportal.bioontology.org/ontologies/BPT) (search by pathway name). DIPG models and figures can be viewed as pathways by links in the file “Supplemental Figures” which listed supplemental figures described in the paper.

Supplemental file “1_PNOC003 gene mutation frequency” contains 9,296 genes with mutations according to WGS.

Supplemental file “2_DE_PNOC003vsGSE120046” – describes differential gene expression in DIPG tumor comparing with GSE120046.

Supplemental file “3_PNOC003 TP53 mutations” – lists of TP53 gene mutations for every patient in the DIPG cohort analyzed in this report. Chromosome position is given for human genome assembly GRCh38.

Supplemental file “4_Regulators of gene expression” – expression regulators with SNEA p-values &lt; 0.05 calculated based on all DIPG samples.

Supplemental file “5_Pathways enriched with regulators (922)” – describes pathways enriched with activity regulators of gene expression in DIPG tumor that have GSEA p-values less than 0.05.

Supplemental file “6_Cells enriched with regulators (922)” – describes cells enriched with regulators of gene expression in DIPG tumor that have GSEA p-values smaller than 0.05.

Supplemental file “7_EZH2 predicted expression targets” – 140 genes that may be hypomethylated and over- expressed in DIPG samples as a result of low EZH2 activity predicted from Elsevier Knowledge Graph.

Supplemental file “8_TP53 predicted expression targets” – 81 genes that may change their expression because of mutations in TP53 predicted from Elsevier Knowledge Graph.

Supplemental file “9_Drugs by SNEA” – 200 drugs targeting regulators of expression of genes in DIPG with best SNEA p-values.

Supplemental file “10_Drugs for mutant TP53” – a list of 585 drugs reported to inhibit mutant TP53 obtained using Elsevier Natural Language Processing technology.

## Data Availability

All data produced in the present work are contained or referred in the manuscript

https://www.researchgate.net/publication/360212757_DIPG_disease_model_Supplemental_materials

## Acknowledgments

We thank Dr. Javad Nazarian for useful discussion and comments that greatly improved the manuscript.

